# Accelerated MRI using intelligent protocolling and subject-specific denoising applied to Alzheimer’s disease imaging

**DOI:** 10.1101/2022.10.24.22281473

**Authors:** Keerthi Sravan Ravi, Gautham Nandakumar, Nikita Thomas, Mason Lim, Enlin Qian, Marina Manso Jimeno, Pavan Poojar, Zhezhen Jin, Patrick Quarterman, Girish Srinivasan, Maggie Fung, John Thomas Vaughan, Sairam Geethanath, the Alzheimer’s Disease Neuroimaging Initiative

## Abstract

Magnetic Resonance Imaging (MRI) is expensive and time-consuming. Protocol optimization to accelerate MRI requires local expertise since each MR sequence involves multiple configurable parameters that need optimization for contrast, acquisition time, and signal-to-noise ratio (SNR). The availability and access to technical training are limited in under-served regions, resulting in a scarcity of local expertise required to operate the hardware and perform MR examinations. Along with other cultural and temporal constraints, these factors contribute to the highly inefficient utilization of MRI services diminishing their clinical value. In this work, we extend our previous effort and demonstrate accelerated MRI via intelligent protocolling of the modified brain screen protocol, referred to as the Gold Standard (GS) protocol. We leverage deep learning-based contrast-specific image-denoising to improve the image quality of data acquired using the accelerated protocol. Since the SNR of MR acquisitions depends on the volume of the object being imaged, we demonstrate subject-specific (SS) image-denoising. Utilizing the accelerated protocol resulted in a 1.94x gain in imaging throughput over the GS protocol. The minimum /maximum PSNR gains (measured in dB) were 1.18/11.68 and 1.04/13.15, from the baseline and SS image-denoising models, respectively.

Alzheimer’s Disease (AD) accounts for up to 60-80% of dementia cases and a global trend of longer lifespans has resulted in an increase in the prevalence of dementia/AD. Therefore, an accurate differential diagnosis of AD is crucial to determine the right course of treatment. The GS protocol constitutes 44.44% of the comprehensive AD imaging protocol defined by the European Prevention of Alzheimer’s Disease project. Therefore, we also demonstrate the potential for AD-imaging via automated volumetry of relevant brain anatomies whose atrophies have been shown to be reliable indicators of the onset of the disease. The volumetric measurements of the hippocampus and amygdala from the GS and accelerated protocols were in excellent agreement, as measured by the intra-class correlation coefficient.

In conclusion, accelerated brain imaging with the potential for AD imaging was demonstrated, and image quality was recovered post-acquisition using DL-based image denoising models.

## I. Introduction

Although Magnetic Resonance Imaging (MRI) is a powerful imaging modality to obtain valuable information about the brain structure anatomy via the acquisition of high-resolution images, multiple challenges are involved with MR imaging. It is expensive and time-consuming, and subjects with MR-unsafe materials (such as medical device implants, prostheses, etc.) are not eligible for MR imaging. Considerable research efforts have been directed toward accelerating acquisition times by exploiting the temporal or spatiotemporal redundancies in the images (Tsao and Kozerke, 2012). However, protocol optimization to accelerate MRI requires local MR expertise. Each sequence involves multiple configurable parameters that need optimization for contrast, acquisition time, and signal-to-noise ratio (SNR). A large number of these combinations exist–for example, 29 million for 12 sequences in a protocol (Block, 2018)–and choosing an optimal combination in real-time is difficult. Since the availability and access to technical training are limited in under-served regions (Geethanath and Vaughan, 2019), this results in a scarcity of local expertise required to operate MR hardware and perform MR examinations. These factors, along with other cultural and temporal constraints contribute to the highly inefficient utilization of MRI services diminishing their clinical value (Geethanath and Vaughan, 2019).

This combination of a very high-dimensional optimization space and inadequate local expertise necessitates a data-driven approach to augment the available manpower. Previous works involve machine learning approaches for automated RF pulse design (Shin *et al*., 2020), sequence design (Zhu *et al*., 2018), or even a joint framework for sequence generation and data reconstruction (Walker-Samuel, 2019; Loktyushin *et al*., 2021). We believe that augmenting human expertise by leveraging deep learning (DL) techniques across the MRI pipeline can consistently yield improved MR Value irrespective of where the service is offered or the expertise involved. MR Value is an initiative by the International Society of Magnetic Resonance in Medicine to measure the utility of MRI in the context of constantly evolving healthcare economics (https://www.ismrm.org/the-mr-value-initiative-phase-1/). We based our prior work on this premise and demonstrated preliminary results from MR Value-driven Autonomous MRI, dubbed AMRI <u>(Ravi et al. ; Ravi and Geethanath 2020)</u>.

Dementia affected 50 million people worldwide in 2018, with an estimated economic impact of US$ 1 trillion a year (Patterson, 2018; Banerjee *et al*., 2020). Alzheimer’s Disease (AD) accounts for up to 60-80% of dementia cases and potentially begins upto 20 years before the first symptoms emerge (Bateman *et al*., 2012). A global trend of longer lifespans has resulted in an increase in the prevalence of dementia/AD (Silva-Spínola *et al*., 2022). An accurate differential diagnosis of AD is crucial to determine the right course of treatment (Vernooij and van Buchem, 2020). Traditionally, structural neuroimaging is used to exclude treatable and reversible causes of dementia such as brain tumors, subdural hematomas, cerebral infarcts, or hemorrhages (Falahati *et al*., 2015). However, studies have demonstrated that atrophy of the hippocampus and amygdala volume are reliable indicators of progression from pre-dementia to AD (Simmons *et al*., 2011). These imaging biomarkers, or image-derived phenotypes (IDP), can be obtained from MRI. The European Prevention of Alzheimer’s Disease (EPAD, https://ep-ad.org/open-access-data/overview) prescribes four core and five advanced sequences for AD imaging. The core sequences are 3D T_1-_weighted (T_1_w), 3D fluid-attenuated inversion recovery (FLAIR), 2D T_2_-weighted (T_2_w), and 2D T_2_*-weighted (T_2_*w). The advanced sequences are 3D T_2_*w, 3D susceptibility weighted imaging (SWI), diffusion-weighted imaging (DWI) or dMRI, resting-state functional MRI, and arterial spin labeling. Furthermore, (Mehan *et al*., 2014) reported on the adequacy of a four-sequence protocol consisting of an axial T_1_w, axial T_2_w FLAIR, axial DWI, and axial SWI images to evaluate new patients with neurological complaints. In this work, we extend our previous effort and demonstrate accelerated MRI via intelligent protocolling of the modified brain screen protocol (dubbed Gold Standard, GS) employed at our institution. We leverage deep learning-based image denoising to improve the image quality of data acquired using the accelerated protocol. The GS protocol consisted of the following sequences: sagittal 3D T_1_w, axial 2D T_2_w, axial 2D T_2_w FLAIR, axial 2D SWI, axial DWI, and axial 2D T_1_w. Overall, the GS protocol constitutes 44.44% of the comprehensive EPAD imaging protocol and includes all sequences deemed adequate for neuroimaging by Mehan *et al*. Therefore, we also investigate the potential of the accelerated protocol for AD-screening by benchmarking volumetry of the hippocampus and amygdala against measurements from the GS protocol. This volumetry can be used for early detection of atrophy.

### A. Study design

Figure 1 is a brief illustration of the processes involved in this work: intelligent protocolling, MR acquisition, DL-based image denoising, and quantitative evaluation. We first demonstrate accelerating the GS protocol employed at our institution using sequence-specific look-up tables (LUTs, Figure 1A). We use LUTs to optimize for accelerated acquisition durations. We perform four experiments designed to measure improvements in metrics such as (i) median object-masked local SNR (Golshan, Hasanzadeh and Yousefzadeh, 2013); (ii) peak SNR (measured in dB); (iii) variance of the Laplacian, referred to as var-Lap (Pech-Pacheco *et al*., 2000); (iv) multi-scale structural similarity index (MS-SSIM, Wang, Simoncelli and Bovik, 2003); and (v) MR Value. We also acquire data from five healthy subjects (Figure 1B) to test for repeatability. Subsequently, we denoise the acquired data using contrast-specific image-denoising models trained on publicly available brain MRI datasets (Figure 1C). Since the SNR of MR acquisitions depends on the volume of the object being imaged, the SNR values of acquisitions of larger subjects will be higher. We obtain preliminary results that illustrate the variability of SNR with varying whole brain volumes and base our rationale for subject-specific (SS) image denoising on this premise. Subsequently, we demonstrate SS-denoising (Figure 1D). Finally, we perform quantitative evaluation using a combination of automated and manual methods.

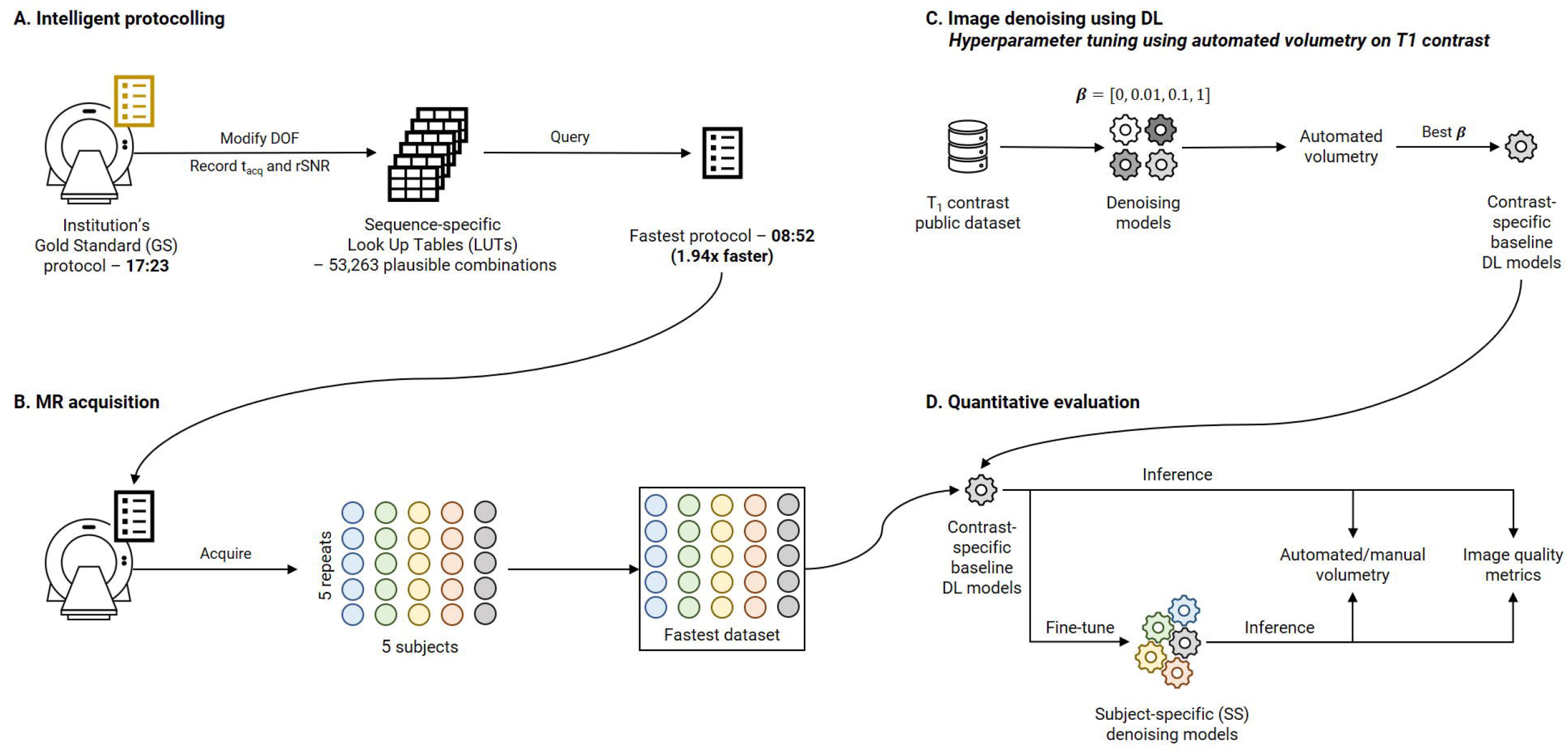

To the best of our knowledge, this work, dubbed Intelligent Protocolling for AMRI (AMRI-IP), is the first framework to integrate the entire pipeline of AD imaging with a particular focus on accelerating MR acquisitions. AMRI-IP provides end-to-end control of acquisition, reconstruction, image processing, and analysis. The flexible and extensible LUT approach can be utilized to accelerate a single sequence, or an entire protocol, with complete control over the optimization criterion. The contrast-specific image-denoising models are inherently generalizable by training on corresponding publicly available datasets. The SS image-denoising models enable tailored image quality recovery for each subject. Finally, the automated quantitative evaluation method enables the measurement of AD-specific IDPs such as hippocampal and amygdala volumes.

In brief, we claim that employing the accelerated protocol yields improvements in MR Value, and achieves comparable image quality to GS as per quantitative evaluation. Section II.D. maps the four experiments to the claims of this work.

## II. Methods and materials

This section is organized as follows. Section II.A. describes intelligent protocolling – accelerating the routine brain screen protocol employed at our institution by consulting a LUT. Section II.B. presents the development of deep learning models to achieve SS denoising and the explainability of the models. The four experiments that were performed to investigate the hypotheses are detailed in Section II.C., and the image-derived phenotypes (IDPs) relevant to AD that were used in performance evaluation are described in Section II.D. Section II.E. discusses the quantitative evaluation methods performed to validate the hypotheses, and finally, Section II.F. describes visualizing intermediate filter outputs for explainable AI.

### A. Intelligent protocolling using look-up tables

Table 1 lists the seven GS sequences and their corresponding acquisition parameters and durations. The cumulative acquisition time was **17:23** (minutes:seconds), as per the vendor console’s user interface (UI). An experienced clinical application specialist was consulted to collate a list of acquisition parameters that could be varied without compromising image contrast for each sequence in the GS protocol. These acquisition parameters were referred to as degrees of freedom (DOF), also listed in Table 1. Exhaustive combinations of these DOF or a hundred randomly chosen combinations, whichever was smaller, were entered into the vendor console’s UI. For each combination (*P*_*acq*_), the acquisition time (*t*_*acq*_) and relative signal-to-noise ratio (rSNR) value were recorded. The *P*_*acq*_, and corresponding *t*_*acq*_ and rSNR values were stored in a LUT. These were searched to obtain the optimal *P*_*acq*_ yielding the lowest *t*_*acq*_. This procedure was repeated for each sequence in the GS protocol. Once the sequence-specific LUTs were constructed, they were consulted to derive sequence-specific optimal *P*_*acq*_ to derive the Fastest protocol. The search procedure is described as follows, applicable to each sequence individually:

#### 1. Compute percentage time allocated

First, the minimum time percentage value (*y*_*1*_) was computed as the ratio of the shortest sequence acquisition time to the shortest protocol acquisition time (*x*_*1*_). Similarly, the maximum time percentage value (*y*_*2*_) was computed from the longest protocol acquisition time (*x*_*2*_). Now, for an imposed protocol acquisition time constraint (*T*_*acq*_), the percentage time allocated (%TA) to a sequence was derived from the straight line fitting the minimum and maximum time percentage values, as described by equation 1:

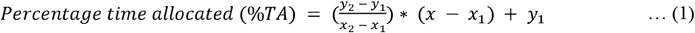

#### 2. Compute weighted rank

The time allocated in seconds for this sequence (*t*_*acq*_) was derived from the %TA value. The LUT was filtered by discarding DOF combinations whose acquisition times exceeded *t*_*acq*_. Of the remaining combinations, weighted differences of rSNR and DOF values with the corresponding default values from the GS protocol were computed. Higher weights were assigned to DOF values contributing more significantly to the image contrast (Supplementary Table 1). Finally, these weighted differences were summed to obtain a rank for each DOF combination, and the resulting LUT was sorted in ascending order of this rank value. Thus, the combination achieving the lowest rank value had the smallest difference in those DOF values which most significantly contributed to the image contrast.

#### 3. Obtain optimal combination

For each time constraint, the combination with the lowest rank was chosen as the optimal set of acquisition parameters.

This process was repeated with lower imposed *T*_*acq*_ in each iteration until an optimal *P*_*acq*_ could not be obtained for every sequence in the GS protocol. In this way, the Fastest protocol was derived by consulting the sequence-specific LUTs. Data acquired utilizing the GS protocol were referred to as the GS dataset. Data acquired from the Fastest protocol for Experiments 1, 2 and 4 (see Section II.D.) were referred to as the Fastest dataset, collectively. The SWI sequence in the GS protocol was replaced by a T_2_*w sequence in the Fastest protocol. The experienced clinical application specialist’s express protocol was dubbed the Expert Express (EE) protocol. Data was also acquired from this protocol for comparison (refer Experiment 2 in Section II.D.), referred to as the EE dataset.

### B. Image denoising using deep learning

Two popular image denoising approaches are to directly predict the denoised image or to obtain the denoised image as the residual of the input noisy image and the predicted noise. We adopt a ResNet-inspired network architecture (demonstrated to improve training performance and stability, (He *et al*., 2016)) to directly predict the denoised output. Individual contrast-specific denoising models were trained on pairs of noisy-denoised images from publicly available brain MRI datasets (see below). Finally, SS denoising was performed by fine-tuning the models on pairs of noisy-denoised images from the prospectively acquired Fastest dataset.

#### Datasets, forward simulation, and data splits

Publicly available datasets were utilized to train the contrast-specific image denoising models. T_1_ and T_2_ contrasts: IXI dataset (https://brain-development.org/ixi-dataset/); T_2_*: ADNI 3 (https://adni.loni.usc.edu); T_2_ FLAIR: MSSEG-2 (Commowick, Cervenansky and Cotton, 2021) and DWI: AOMICID-1000 (Snoek *et al*., 2021). Wherever applicable, datasets were filtered to retain only the 3T data. Only the central 50% slices were utilized, and the remaining slices were discarded to avoid either unwanted anatomy or pure background noise. Supplementary Figure 1 presents the search criteria that were utilized to filter the ADNI 3 dataset for relevant results. For DWI, only the b0 images were utilized from the AOMICID-1000 dataset. All these datasets were assumed to be free of MR image artifacts, referred to as ‘clean images’. Figure 2 presents the forward modeling process to generate noise-corrupted data (‘noisy images’), described here as follows. First, the object-masked local SNR maps were computed on all acquisitions of an arbitrarily chosen subject from the Fastest dataset (see Experiment 4, Section II. D.). Object masking was based on the technique in (Jenkinson, 2003) and resulted in all background values being set to zero. All remaining non-zero values were considered to belong to the samples, which were non-skull-stripped brain images. The local SNR maps were computed based on the method reported in Golshan, Hasanzadeh and Yousefzadeh, 2013. The volume yielding the lowest median SNR was the ‘noisiest acquisition’ (Figure 2A). Next, motivated by work in (Geethanath, Poojar and Ravi, 2021; Qian *et al*., 2022), native noise values were extracted from this noisiest acquisition. These noise values were collaged to form a native noise block (Figure 2B). This was randomly sampled to obtain noise values, which were scaled and added to the clean images to obtain noisy images (Figure 2C). The scaling factor was determined using an iterative approach. A volume was chosen at random from the public dataset. Starting with an initial value of 1.0 (corresponding to no scaling), the scaling factor was increased by 0.5 in each iteration until the median object-masked local SNR of the corrupted volume was lesser than that of the noisiest acquisition. We chose median over mean as the guiding measure since it was less affected by skewed distributions. An 85%-10%-5% subject-wise split was performed to form the train, validation, and test sets.

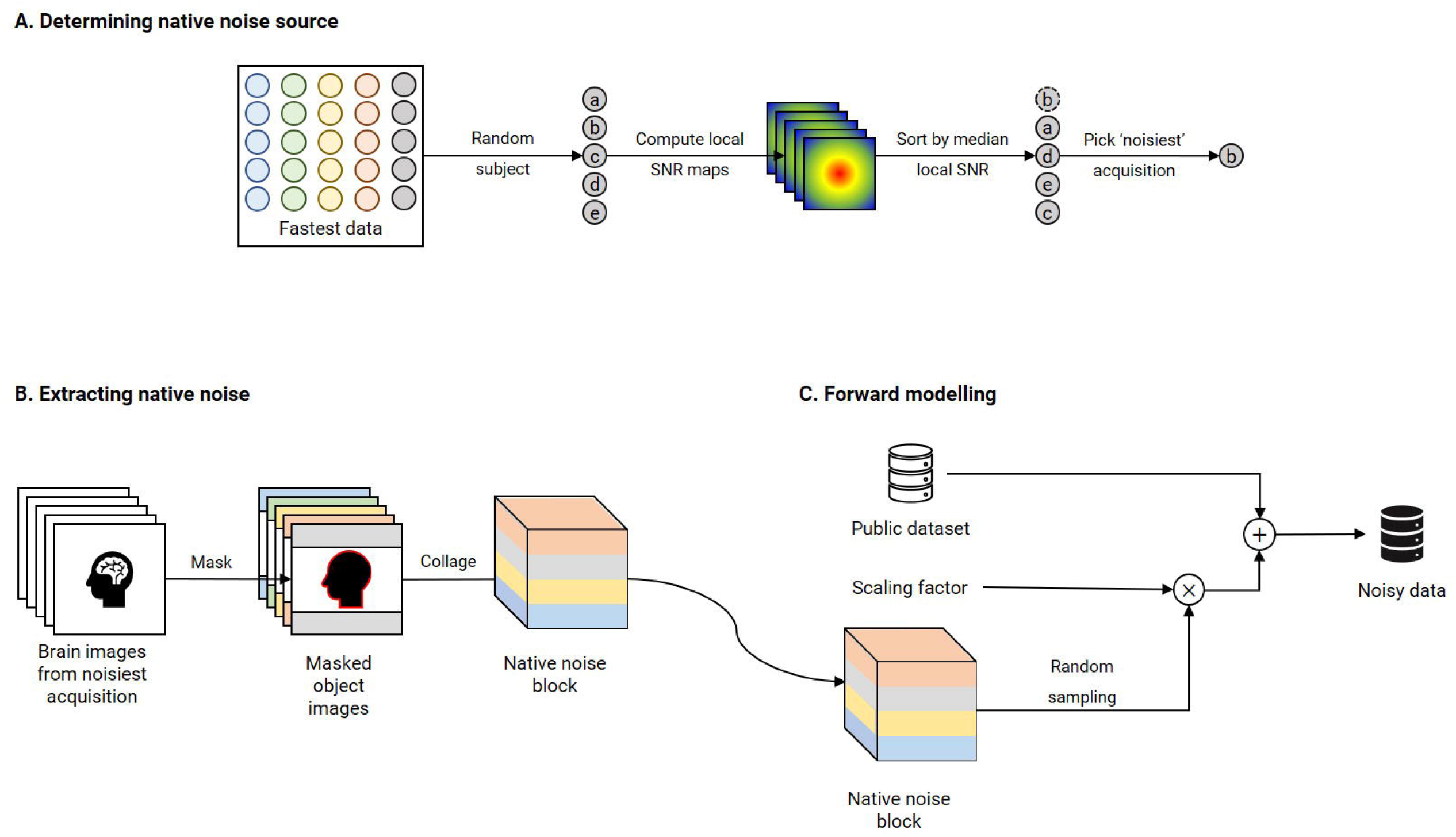

#### Network architectures

Figure 3A is an illustration of the network architecture common to all contrast-specific image-denoising models. To predispose the network to learn denoising filters whilst being anatomy agnostic, we adopted a patch-wise approach in this work. Overlapping patches of size 64 × 64 were input to the network. Thirteen ResBlocks leveraged skip connections to improve training performance (He *et al*., 2016). Each ResBlock consisted of two ReLU-activated (Nair and Hinton, 2010) 3 × 3 2D convolution layers. In case of a mismatch in the number of filters between the previous and current ResBlocks (*N*_*1*_, *N*_*2*_), the skip connection included a 1×1 2D convolution with *N*_*2*_ filters. Otherwise, the skip connection was an identity operation. Additionally, an identity skip connection was used to add the input data to the pre-final layer in the overall network. The final layer was a 3x 3 2D convolution layer with 1 filter. All 2D convolution layers were ReLU-activated, and all development, training, and testing were performed using Keras 2.6/TensorFlow 2.6.2 (Abadi *et al*., 2016; M. Abadi *et al*., 2016) libraries.

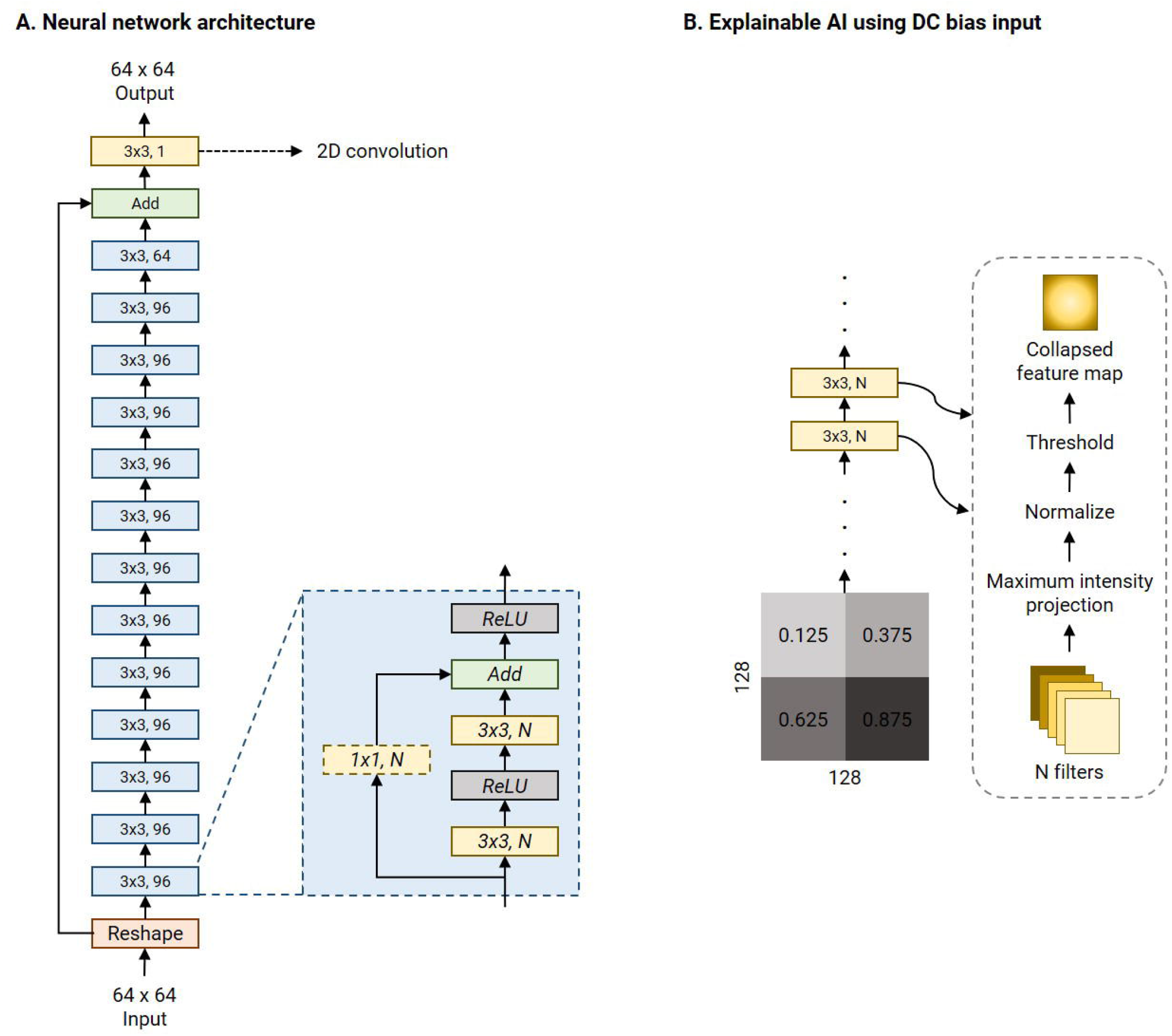

#### Loss functions

Zhang et al. report the superiority of their mixed loss (Mix-L) function for image denoising, among other image quality restoration applications (Zhao *et al*., 2017). This loss is a weighted sum of *l*_1_ and MS-SSIM losses:

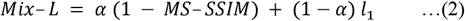

… where *α* was set to 0.84. We modified Mix-L to incorporate a data-consistency term with the measured data in the Fourier domain, referred to as Mix-L+FTD:

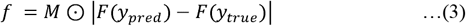

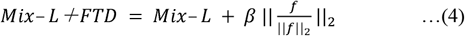

… where *F* was the 2D Fourier Transform and *M* was a mask to only retain the central crop of the k-space of size 16*x*16. The ⊙ operator represented the Hadamard product and *β* determined the trade-off between the denoising and data-consistency errors. We investigated *β* = [0, 0.01, 0.1,1] in our experiments. To determine the best *β*, the RMSEs of the volumetric measures (RMSE_vol_) were computed using an automated tool on T_1_ denoised outputs. The *β* yielding the lowest mean RMSE_vol_ was chosen to train the denoising models for the remaining contrasts. Section II.C. describes the automated T_1_ volumetry tool and computing RMSE_vol_ in detail.

#### Training

All contrast-specific denoising models were trained for 100 epochs with a batch size of 256. The Adam optimizer (Kingma and Ba, 2014) was utilized to minimize the Mix-L+FTD loss with the optimal *β*, determined as stated above. During training, every input slice was cropped to a 64 × 64 patch. The bounds for the random crop were manually determined by examining the corresponding public dataset such that the random crops would mostly include brain anatomy. A callback was utilized to save the model achieving the lowest validation loss (corresponding to ‘best performance’). At the end of the training process, this model was chosen as the best model for evaluation, including to determine the optimal *β*.

#### Subject-specific (SS) denoising

SS median local SNRs were computed on masked brain volumes from the Fastest dataset to verify the premise of SS denoising. The values were computed only on the central 50% of the slices. Next, the same noise scaling factors were utilized to corrupt each subject’s noisiest acquisition from the Fastest dataset with native noise. This data was used to fine-tune the baseline denoising models to achieve SS denoising. This approach posed SS denoising as a self-supervised learning problem, mimicking the noisy-as-clean method demonstrated by (Xu *et al*., 2020). The initial learning rate of the Adam optimizer was reduced to avoid large modifications to the weights which would otherwise harm the learned representations (Table 2).

### C. Experiments

We performed four experiments to investigate hypotheses regarding the throughput of the Fastest protocol, and the image quality of the Fastest dataset. Supplementary Table 2 lists the experiments performed, the protocols executed, numbers of healthy volunteers imaged, the corresponding claims and hypotheses investigated, and their respective evaluation criteria. In total, 31 brain volumes were acquired from five volunteers across the four experiments. The data acquired from the GS and Fastest protocols are referred to as the GS and Fastest datasets, respectively.

#### Experiment 1 (E1) – Throughput

The goal of experiment one was to investigate if the Fastest protocol obtained from the LUT would yield an improvement in throughput. One volunteer was imaged using the GS and Fastest protocols, and a video recording of the entire imaging session was captured. Throughput was computed as the ratio of the table time measurement of the Fastest protocol to that of the GS protocol. In this work, table time is defined as the duration between the scanner bed reaching the center of the bore at the start of the imaging and the scanner bed returning to the home position. We also determined the MR Values of the GS and Fastest protocols, calculated as the ratio of the cumulative median object-masked local SNR values across all contrasts to the protocol’s acquisition duration, T_acq_:

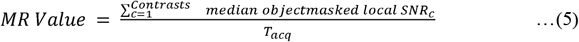

The object-masked local SNR maps were computed on the central 50% slices across all sequences in each protocol.

#### E2 – Image quality

Experiment two quantitatively compared the image quality of the GS, EE, and Fastest datasets using the following metrics: median object-masked local SNR and var-Lap. PSNR and SSIM were not used since they are not reference-less metrics.

#### E3 – SNR recovery

We investigated the feasibility of employing the Fastest protocol. It involved utilizing the image-denoising deep learning models described in Section II.B. to improve image quality. The metrics described in Section II.D. were utilized to determine if denoising the Fastest dataset achieved comparable quality to the GS dataset.

#### E4 – Repeatability

A repeatability test to demonstrate the consistency of the quantitative image quality metrics was performed. The GS and Fastest protocols were employed to acquire data from five subjects over five repeats. Automated and manual volumetry were performed on the acquired data as described in Section II.D.

### D. Quantitative evaluation

<u>Thomas et al., 2020</u> demonstrated an end-to-end pipeline for fully automated mental health screening. The authors leveraged a DL model to segment the various subgroups. Further development on the previous work includes a second DL model to segment the brain tissues (white matter, gray matter, cerebrospinal fluid). This second DL model was based on the nnUnet (Isensee et al., 2021), and an evaluation of its performance is presented in Supplementary File 1.

We leveraged this tool to perform automated volumetry to measure the performance of the denoising models. HTML reports were generated containing volumetric measures of 27 brain subregions and 3 brain tissues. These were programmatically extracted and tabulated. RMSE_vol_ was calculated as the mean of RMSEs of each of the volumetric measures. A benign White Matter Hyperintensity (WMH) was identified in data acquired from one subject. The free, open-source, and multi-platform 3D Slicer software (https://www.slicer.org/, Fedorov et al., 2012) was used to perform manual volumetry of this WMH on the T_2_, T_2_*, T_2_ FLAIR and DWI data by four different raters with 3-8 years of MR experience. All volumetries were performed on data acquired for E4 of the Fastest dataset.

Additionally, a set of image quality metrics were also computed to evaluate the denoising models. These were: median object-masked local SNR, PSNR, MS-SSIM, var-Lap and MR Value. While local SNR, PSNR, and MS-SSIM metrics are commonly used to measure image quality, we obtain var-Lap values to measure the amount of blurring. We included this metric in our evaluations since blurring negatively affected the automated volumetry on T_1_ (preliminary experiments not reported in this work).

### E. Visualizing learned filters for explainable AI

A 256 × 256 collage of four panels was assembled to investigate the features learned by the filters of the 2D convolution layers. Each of the four 64 × 64 panels was made up of a single intensity from the following values: [1.25, 3.75, 6.25, 8.75] × 10^−1^. This collage was corrupted with native noise from an arbitrarily chosen subject’s T_1_w acquisition. Subsequently, it was denoised using the baseline denoising model for the T_1_ contrast. The filter outputs of each 2D convolutional layer were obtained, and maximum intensity projection was performed to achieve dimensionality reduction. Therefore, this collapsed N filter outputs into a single map. This was normalized to lie in the range [0, 1.0]. Finally, this collapsed feature map was hard thresholded to only retain values > 0.75. Figure 3B briefly illustrates this procedure.

### F. Statistical analysis

The intra-class correlation coefficient (ICC) was calculated based on the analysis of variance (ANOVA) with repeated measures to assess the agreement of the volumetric measures amongst the GS, denoised baseline, and denoised SS methods. The ICC values greater than 0.9 indicate excellent agreement, values between 0.75 and 0.9 indicate good agreement, and values between 0.5 and 0.75 indicate moderate agreement.

## III. Results

### A. Intelligent protocolling and LUTs

rSNR was assigned the smallest weight when constructing the LUT since the aim of this work was to achieve acceleration by trading-off SNR which could be recovered post-acquisition via deep learning methods. The total duration of the Fastest protocol that was obtained by querying the LUTs was 8:34 (minutes:seconds). This was a 50.71% reduction in acquisition time from the GS protocol, which required 17:23. The EE protocol only required 7:12. It primarily achieved acceleration by employing the 3D T_1_ FLAIR sequence. This is not a true 3D acquisition and only required 0:37 when compared to 2:44 and 1:41 for the 3D T_1_w sequences in the GS and Fastest protocols, respectively. Figure 4 is a collage of one representative slice of an arbitrarily chosen subject, across contrasts. The rows represent the different datasets–GS, EE, Fastest, baseline denoised, and SS denoised. For the Fastest protocol, the sagittal T_1_-MPRAGE sequence was accelerated by increasing the slice-thickness from 1.0 mm in the GS protocol to 1.6 mm. Overall, this resulted in increased signal intensities and decreased variance of noise in the Fastest dataset. Therefore, the median local SNR of GS data was lower than that of the Fastest data, as is expected.

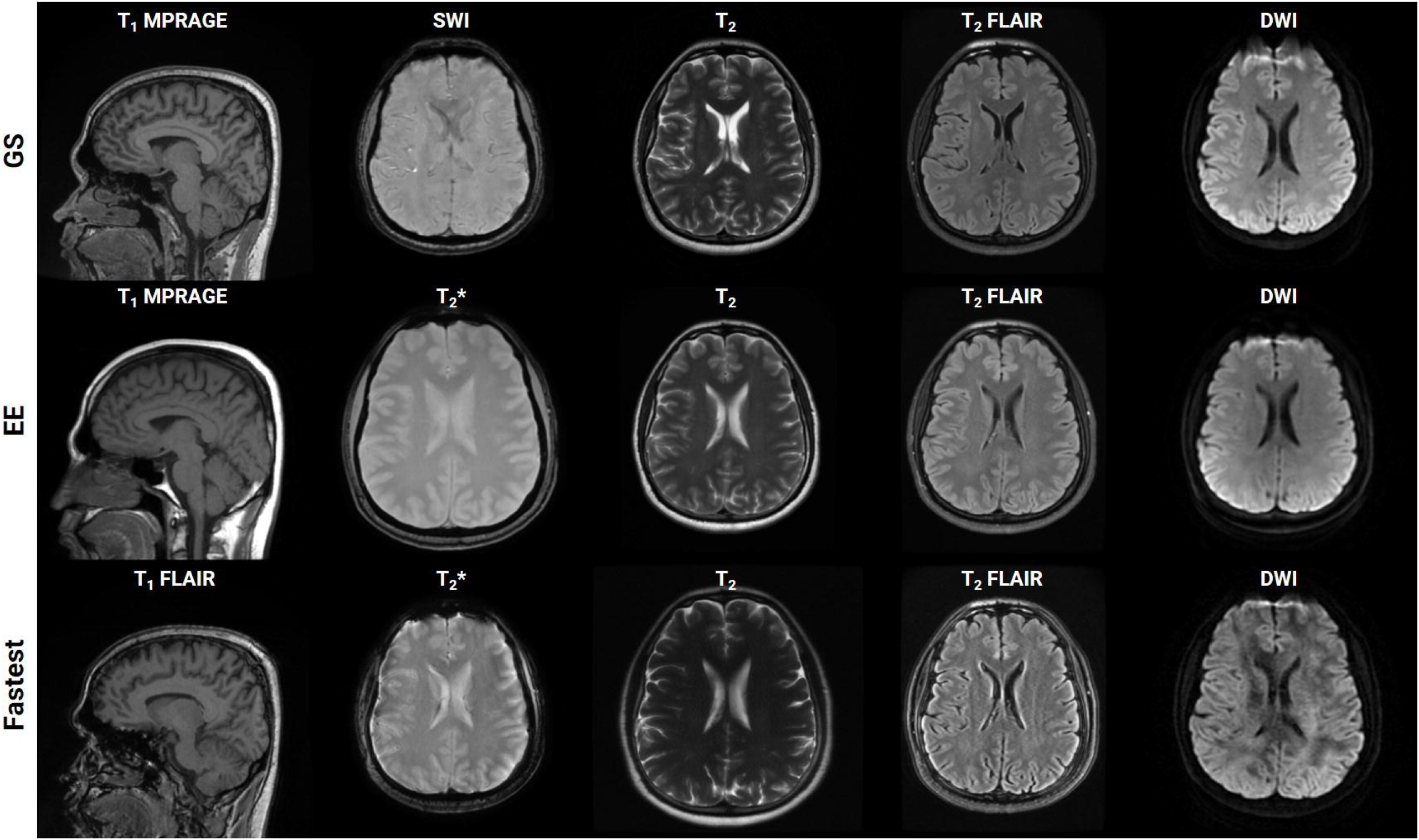

### B. Image denoising using deep learning

#### Datasets and forward-simulation

The T_1_w and T_2_w datasets consisted of 185/13,690 and 185/11,760 volumes/slices, respectively. For T_2_* and T_2_ FLAIR, this resulted in 89/2,188 and 40/6,792 volumes/slices, respectively. Finally, the DWI dataset contained 81/2,430 volumes/slices. The final noise scaling factors determined using an iterative local SNR-guided approach were as follows. T_1_: 1.5; T_2_: 2.0; T_2_*: 1.0, T_2_ FLAIR: 1.0, DWI: 1.5. Supplementary Figure 2 plots the maximum, minimum and mean (dashed line) local SNR values within a region of interest across the GS and forward modeled datasets, for T_1_ contrast. It can be observed that the means of the GS and forward-modeled data are comparable, which validates the iterative local SNR-guided approach to determining the noise scaling factor.

#### Loss functions

Supplementary File 2 presents a tabulation of volumetric measures of denoised data obtained from the automated volumetry tool. It compares the values of data denoised using the models trained on f3 = [0, 0.01, 0.1,1]. The model trained on f3 = 1 achieved the lowest mean RMSE value, and all subsequent models were trained with the same loss formulation.

#### Training

Supplementary Figure 3 shows a plot of training and validation losses as a function of epochs for the baseline denoising models, across contrasts. The corresponding approximate training durations are also listed. Figure 5 presents the corresponding mean changes in the image quality metrics computed on the test sets. In each instance, the model with the lowest validation loss was used. The largest gains in PSNR, MS-SSIM, and var-Lap values are observed on the T_2_ contrast. Since the network architecture was common to all contrast-specific denoising models, this could be attributed to the larger noise scaling factor during the forward modeling process. Consequently, this might have forced the model to learn to denoise much noisier images during training in comparison with the training data from other contrasts. The lowest gains are observed on the T_2_* contrast. This could potentially be because the publicly available T_2_* dataset was corrupted by native noise sourced from an SWI acquisition (GS protocol).

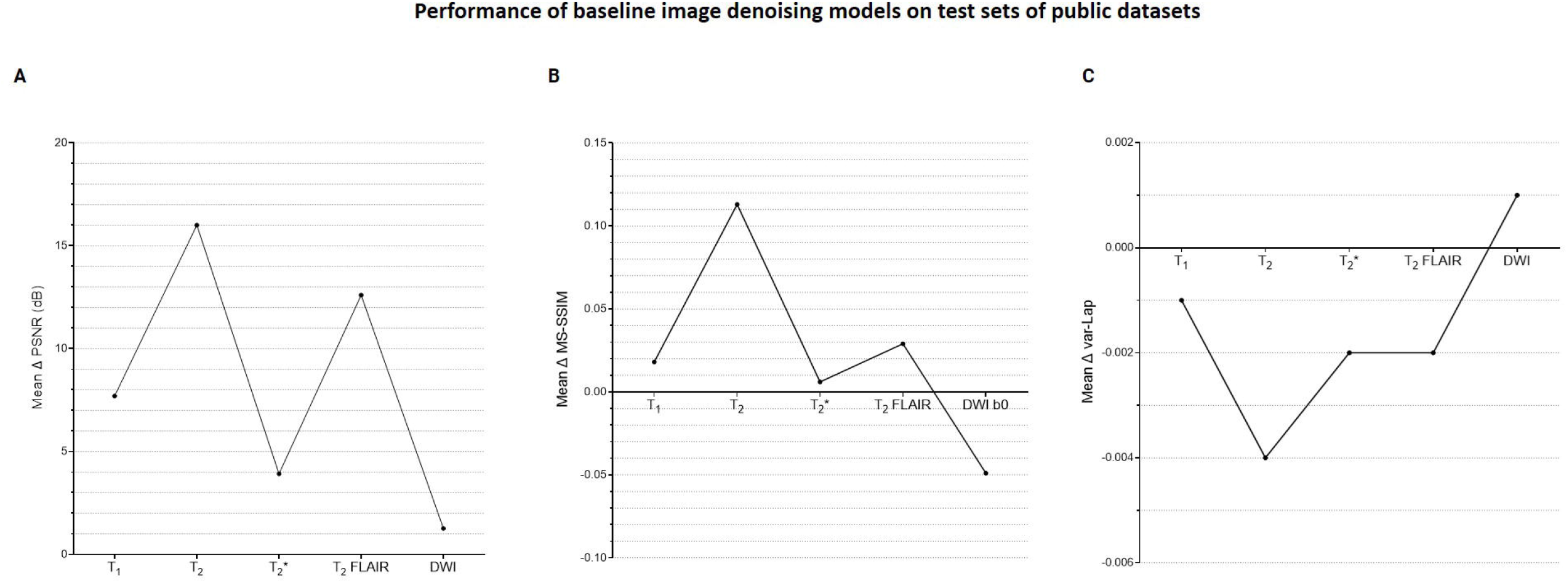

#### SS denoising

Figure 6 plots the subject-specific median local SNR values. The subject-dependent variability of SNR validated our rationale for subject-specific denoising. Figure 7 plots the mean changes in PSNR, MS-SSIM, and var-Lap values across contrasts and subjects. Similar to the baseline denoising models, the largest gains are observed for the T_2_ contrast, and the least improvement is observed for the T_2_* contrast. In addition, T_2_* SS model performed significantly poorer than the baseline model, and therefore the results have not been reported. We suspect this is due to the mismatch in the training and fine-tuning datasets. The ADNI-3 training dataset consisted of T_2_* GRE acquisitions with an echo train length (ETL) of 3. On the other hand, the Fastest protocol utilized an ETL of 1 in the T_2_* GRE acquisitions. We attribute the inherent mismatch in signal between the datasets to the poor fine-tuned performance. Additionally, this potentially indicates that the pre-processing steps in this work are inadequate.

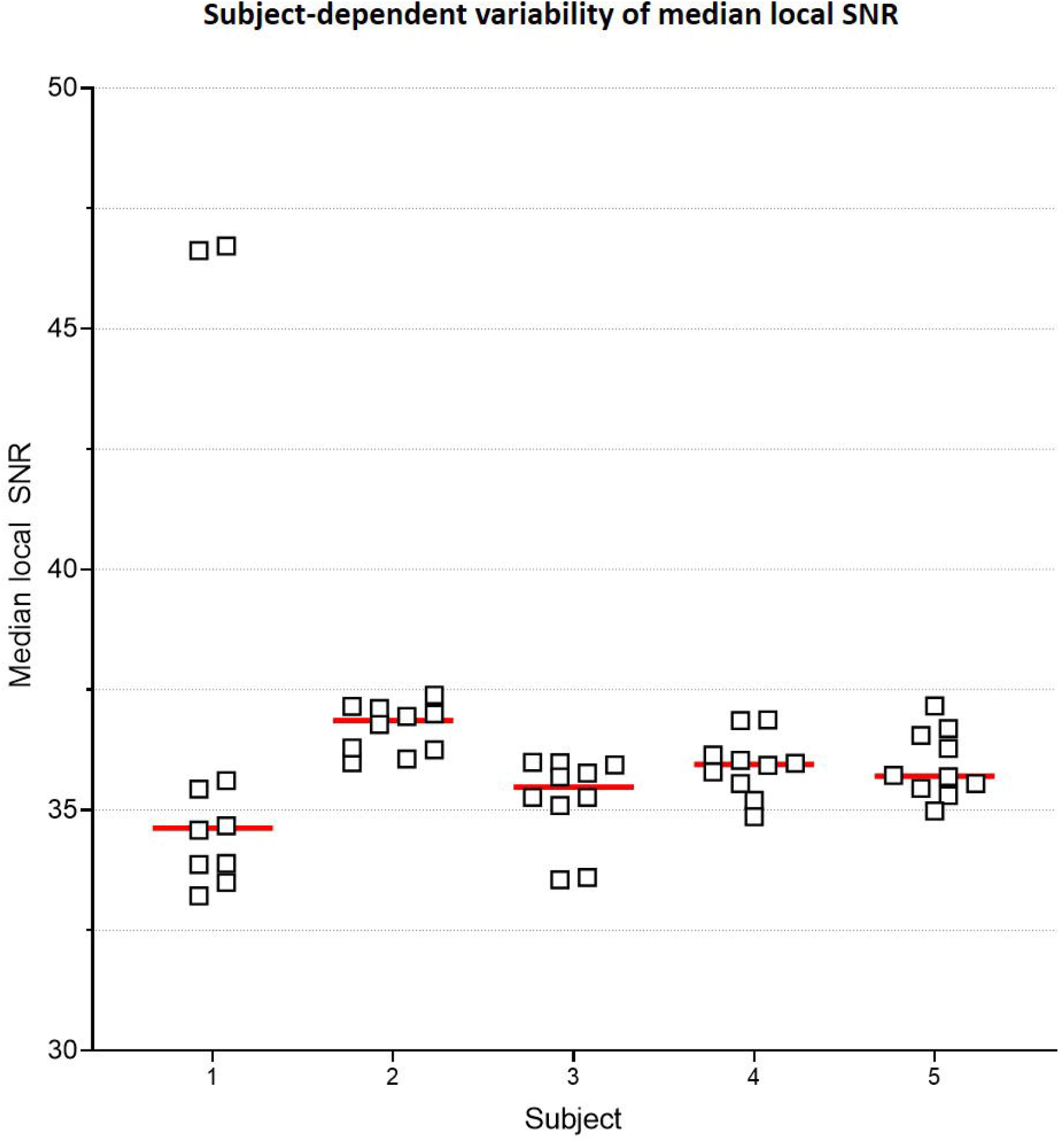

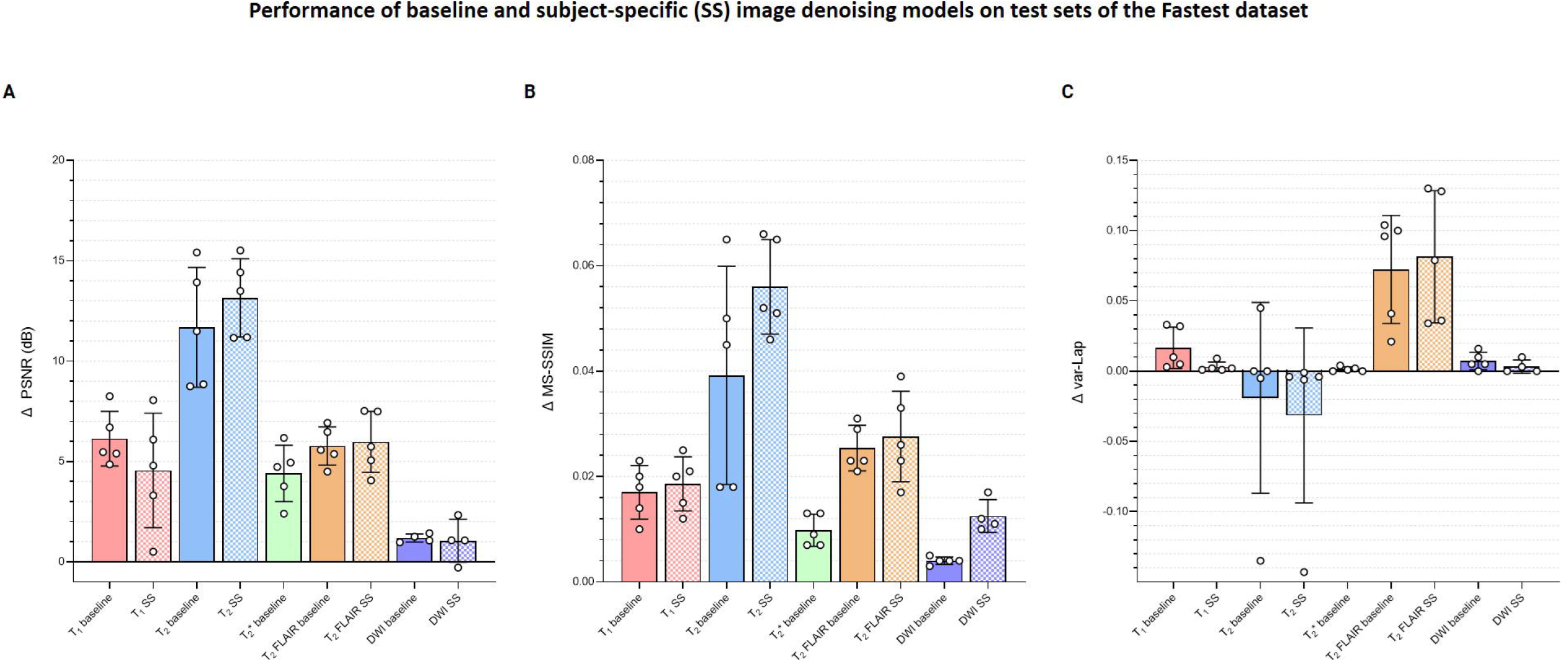

### C. Experiments

#### E1 – Throughput

The cumulative acquisition times for the GS and Fastest protocols as per the vendor UI were 17:23 and 8:34. The practical acquisition times (obtained from the video recording) were 19:12 and 9:52. This discrepancy can be attributed to the time lost during pre-scan calibration and shimming functions. Overall, imaging one healthy volunteer using the Fastest protocol yielded a 1.94x gain in throughput over the GS protocol. Supplementary File 3 presents the timestamps and calculations of durations derived from the video recordings to obtain the final acquisition durations for this experiment. The cumulative median object-masked local SNR values for the GS and Fastest data were 243.354 and 215.767, respectively. Finally, this translates to MR Values of 0.211 and 0.364, respectively. Overall, employing the Fastest protocol resulted in a 72.51% increase in MR Value. In comparison, the corresponding SNR value for EE data was 264.136. Considering a practical acquisition duration of 07:58, this resulted in an MR Value of 0.552.

#### E2 – Image quality

Figure 8 is a bar graph plotting the median object-masked local SNR and var-Lap values across contrasts, for the GS, EE, and Fastest datasets. The mean values are presented at the bottom of the individual bars. For local SNR, similar performance is observed from the axial DWI and axial T_2_ FLAIR sequences, while not for the other sequences. The higher median local SNR values of the T_1_ contrast from the EE protocol can be attributed to the Turbo Spin Echo-based FLAIR sequence. The GS and EE protocols also perform better than the Fastest protocol in the T_2_ sequence, attributed to the longer repetition times. Overall, the EE protocol yields higher local SNR values due to higher slice thickness: 5 mm across all sequences, as opposed to ranges of 1.0–3.6 mm and 1.6–5.0 mm for GS and Fastest protocols, respectively. For var-Lap, comparable performance is observed only in the T_2_ FLAIR contrast. The Fastest protocol performs worse than both GS and EE in T_1_, DWI and T_2_ contrasts.

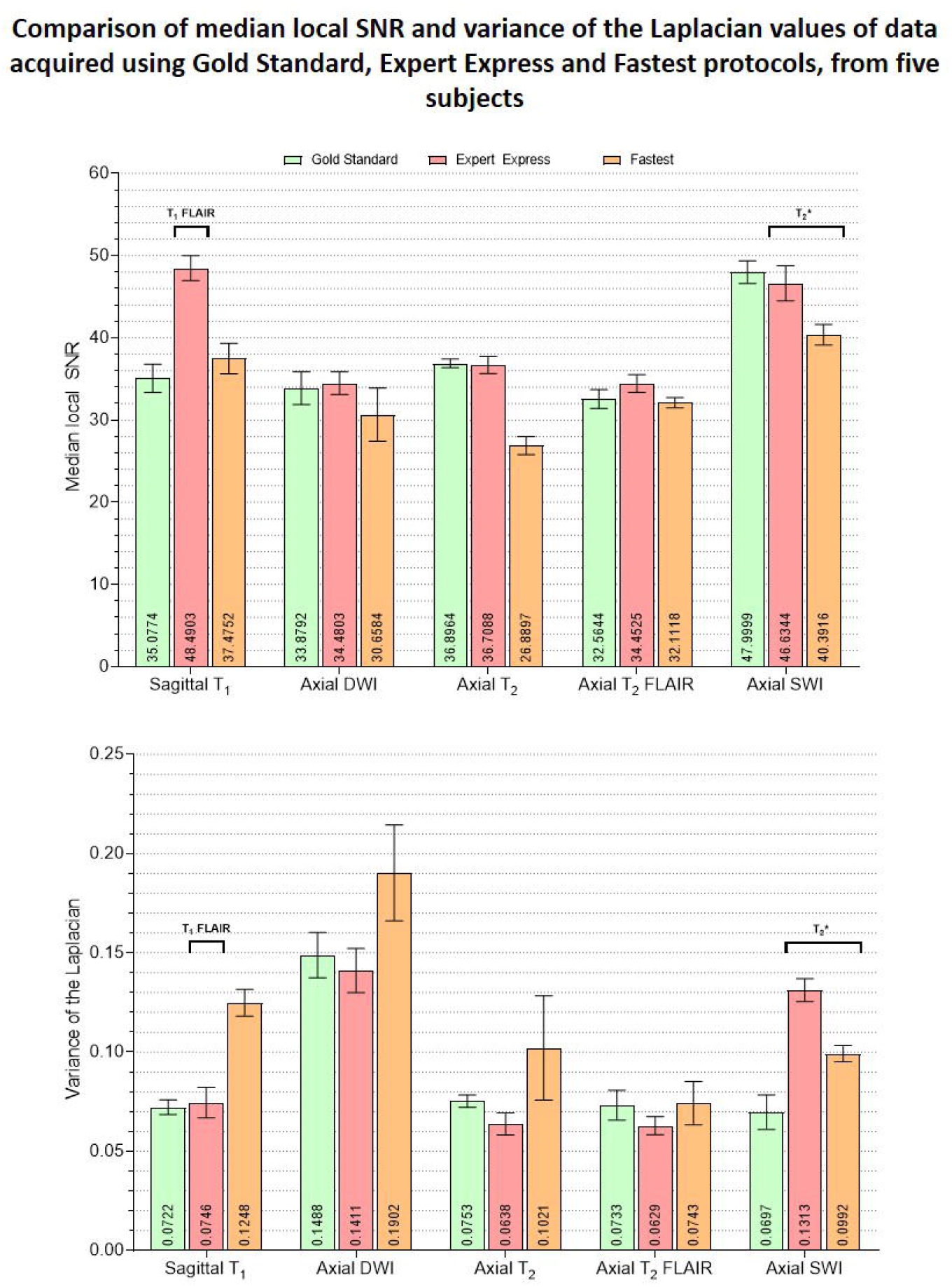

#### E3 – SNR recovery

Figure 6 presents the changes in median object-masked local SNR, PSNR, SSIM, and var-Lap values for the baseline and SS denoising models tested on the Fastest datasets. The solid and checkerboarded bars correspond to the baseline and SS denoising models, respectively. The T_1_ SS denoising model does not improve PSNR over the baseline denoising model, and only modestly improves SSIM. However, it results in a smaller increase in blurriness. For T_2_, the SS denoising model yields larger improvements across all metrics. For T_2_ FLAIR, similar improvements are observed for PSNR and MS-SSIM, along with an undesirable increase in blurriness–indicating the model potentially denoised by primarily high-pass filtering. The T_2_* SS denoising models deteriorated image quality in every instance, and hence their results are not presented.

#### E4 – Repeatability

Figure 9 presents the plots of volumetric measures obtained from the automated tool for T_1_ contrast. The top row plots values of White Matter (WM) and Gray Matter (GM), and the bottom row corresponds to measures of two IDPs for AD–hippocampal and amygdala volumes. For each of these anatomies, a representative slice with the corresponding masks overlaid is illustrated in the figure inset. Figure 10 is a plot of manual volumetric measures for T_2_, T_2_*, T_2_ FLAIR and DWI contrasts.

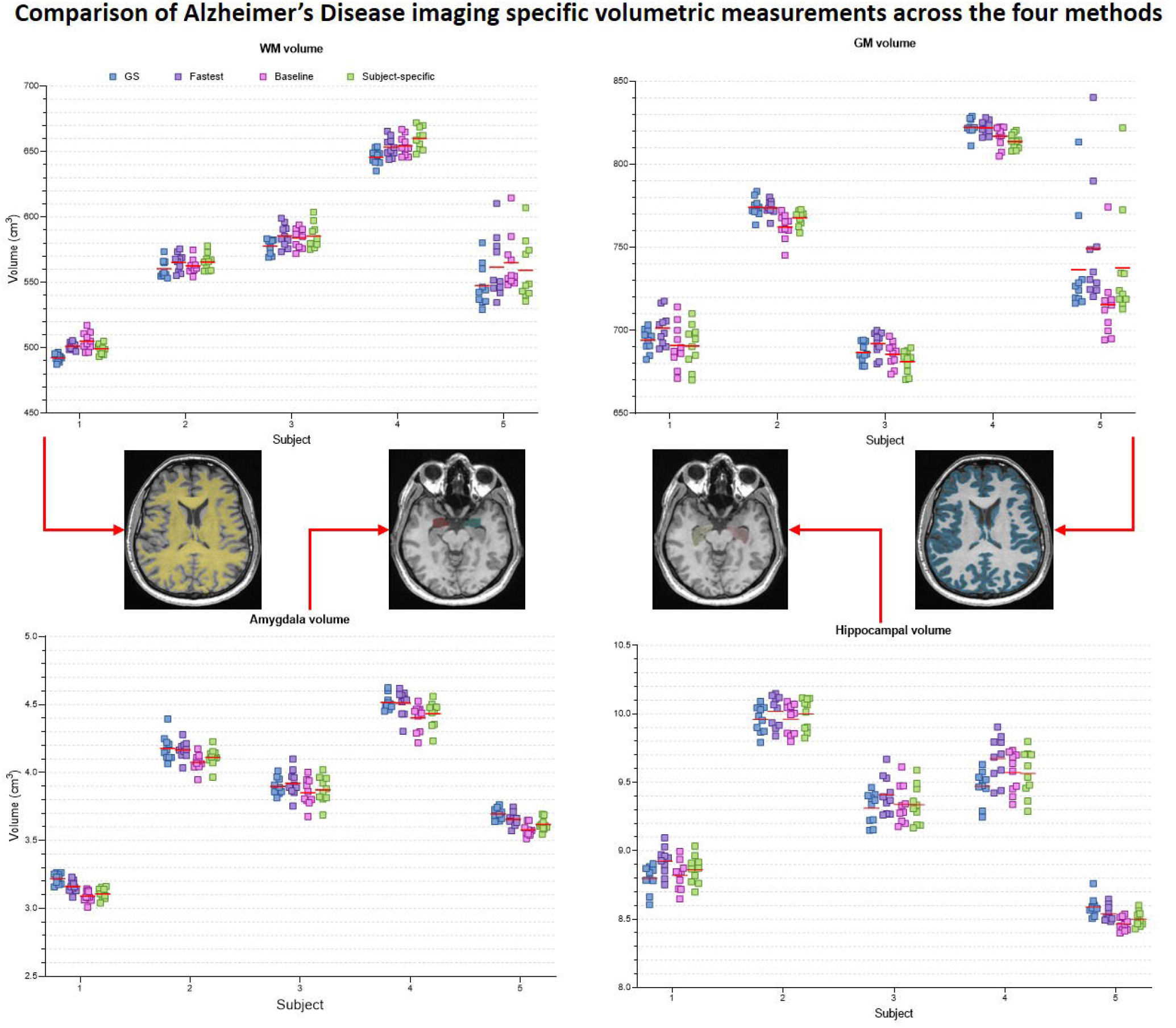

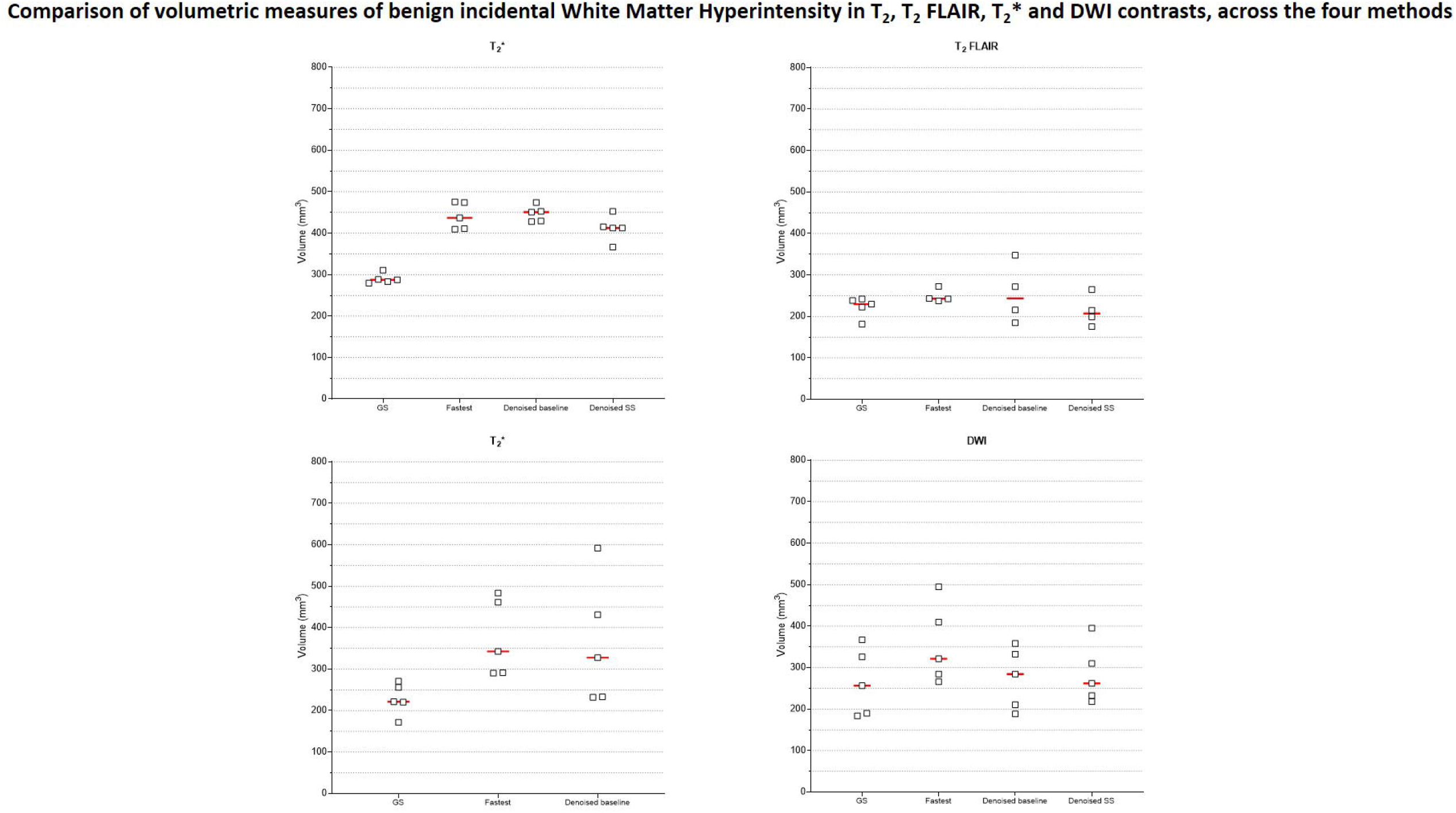

### D. Visualizing learned filters for explainable AI

Figure 11 is a collage of intermediate layer outputs obtained from denoising a DC-biased input using the baseline image denoising model for the T_1_ contrast. The model appears to perform denoising (akin to low-pass filtering) in the earlier layers. Each MaxPool2D layer halves the spatial dimension, leading to reduced resolution in the later layers (refer network architecture in Figure 3A). In these layers, the model appears to be performing low-frequency denoising (high-pass filtering). Overall, no brain-specific anatomy is identifiable across any of the intermediate layer outputs (as desired), potentially attributed to the patch-wise approach adopted in this work.

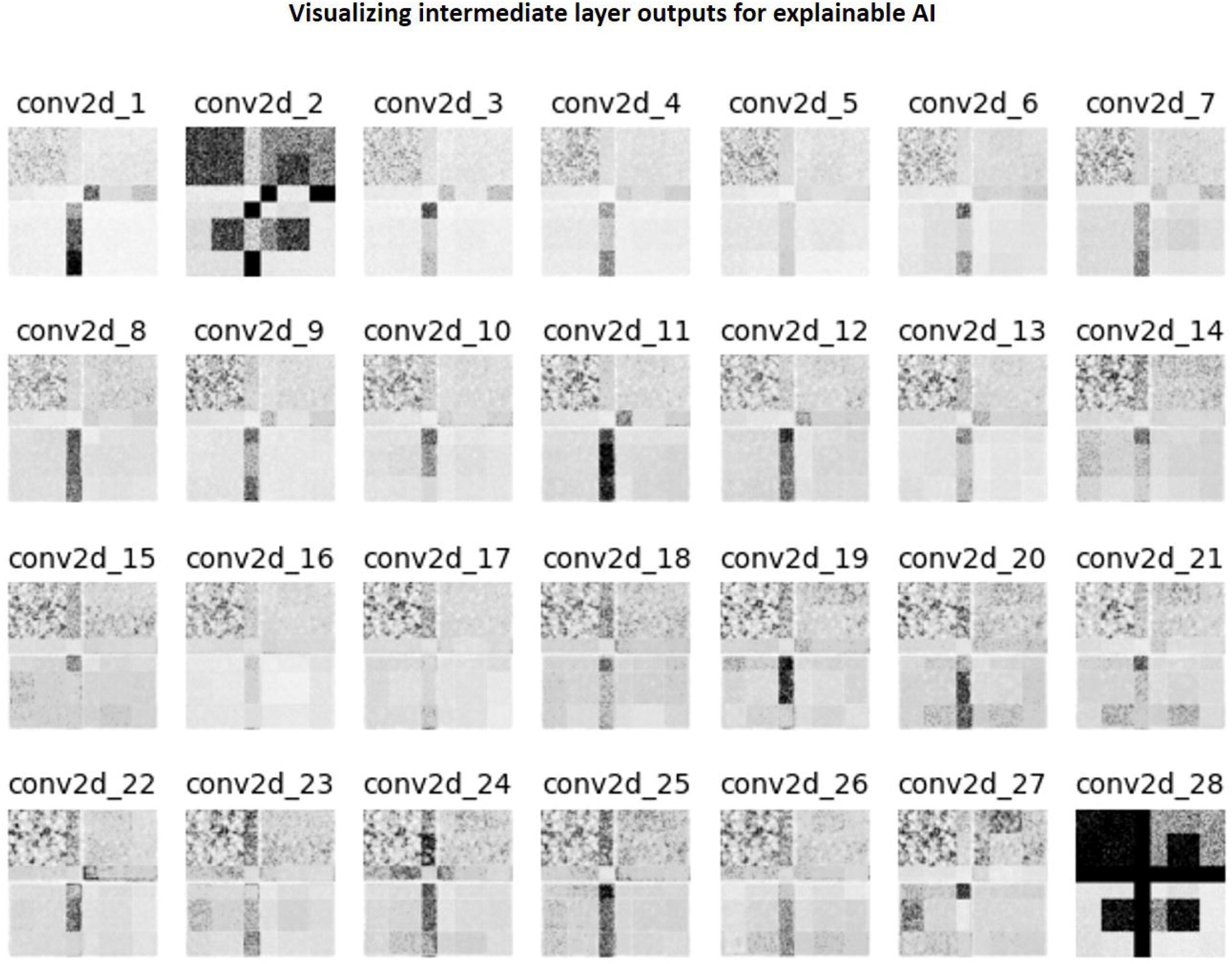

### E. Statistical analysis

Among the four methods for all 30 locations, 27 locations had excellent ICC (>=0.93); 2 had a good ICC (>0.8), 1 had moderate ICC (=0.651). Table 3 lists the individual ICC values for each of the 27 brain subregions and the 3 brain tissues.

## IV. Discussion and conclusion

The LUT search to accelerate the GS protocol was automated. In comparison, designing the EE protocol required human expertise and manual hours. The LUT approach is also scalable– automated recording of acquisition times and rSNR values from the vendor UI for different *P*_*acq*_ can potentially enable the construction of high-dimensional LUTs. Subsequently, high-dimensional constrained search techniques can be explored to arrive at different *P*_*acq*_. Our LUT search formulation also allows optimizing for different criteria. We optimized for shorter acquisition durations whilst trading-off SNR. However, this can easily be modified to any other criteria by suitably modifying the weights described in Supplementary Table 1. Or, the LUT search can involve finding optimal *P*_*acq*_ that satisfies an imposed acquisition time constraint, as demonstrated in our previous work <u>(Ravi et al.)</u>. Furthermore, since domain expertise is involved in setting the weights for the DOF, the LUT search is inherently explainable.

Initially, we trained our image-denoising models for T_1_ and T_2_ contrasts on the Human Connectome Project dataset (HCP, http://www.humanconnectomeproject.org/). Preliminary results (not reported in this work) indicated poor accuracy on the automated volumetry (high RMSE_vol_), although the denoising performance was good. We attributed this to HCP data’s superior image quality–HCP data were acquired on Siemens Prisma 3 T scanners with 80 mT/m gradient strength and 200 T/m/s slew rate. A 3D T_1_ MPRAGE sequence was utilized with isotropic resolution and repetition/echo times = 2530/1.15 ms. Therefore, the iterative local SNR-guided approach resulted in a higher noise scaling factor to degrade the HCP data to match the median local SNR with that of the Fastest dataset. We suspect that the denoising models trained on this data caused excessive blurring, which subsequently affected the automated T_1_ volumetry. Therefore, we chose to proceed with the IXI dataset for T_1_ contrast, and also for T_2_ contrast to potentially mitigate a similar issue.

For T_2_*, the SS denoising models failed to demonstrate better performance than the baseline denoising models. The baseline model was trained on T_2_* data of the ADNI 3 dataset corrupted by native noise extracted from SWI data. However, fine-tuning the baseline model involved training on T_2_* data corrupted by native noise extracted from T_2_* data itself. We suspect this sequence-specific noise distribution could have impacted the training process of the SS models.

### A. Limitations and future work

#### Intelligent protocolling

The T_1_ MPRAGE sequence in the Fastest protocol achieved shorter scan durations due to higher slice thickness (1.6 mm vs 1 mm). Future work could involve exploring the impact of interpolating anisotropic data to achieve isotropic voxel resolutions on the accuracy of automated volumetry (Deoni *et al*., 2022). Although our LUT search formulation was designed to avoid modifying image contrast, the Fastest dataset marginally deviates from the GS dataset’s contrast. For example, this can be observed in the T_2_ FLAIR contrast in Figure 4. Potentially, Virtual Scanner (Tong *et al*., 2019) and its digital twinning capability (Tong, Vaughan and Geethanath, 2021) can be leveraged to design a physics-informed LUT optimization approach.

#### Data distribution

For detecting AD, volumetry from T_1_-MPRAGE sequence is crucial. The denoising models were evaluated on a small and healthy cohort of five volunteers. Their performance on denoising pathological data has not been investigated. While we have demonstrated the value of denoising in improving the accuracy of volumetry, the robustness of the denoising models on out-of-distribution data has not been considered. A thorough evaluation will be required to assess the quality of data acquired from pathological subjects and denoising using our models. Alternatively, the training dataset could include pathological data to improve the models’ generalization capabilities. Datasets which have not undergone extensive preprocessing and/or stringent quality control are valuable during the native noise extraction process of our workflow. Future iterations could involve training on a multi-site, multi-vendor, real-world dataset such as RadImageNet (Mei *et al*., 2022).

#### Evaluation metrics

This work utilizes a combination of referenceless (local SNR, var-Lap) and reference-based (PSNR, SSIM) image quality metrics. The referenceless metrics were borrowed from the broader computer vision community, and might not be ideal for evaluating methods in medical imaging. In particular, since the var-Lap metric is on an arbitrary scale, it does not allow performance comparisons without controlling for the testing dataset. The reference-based metrics inherently require a gold standard (GS), and hence do not lend themselves to evaluation on real-world data which, by nature, do not have reference data.

#### Inference on denoising models

While the patch-wise implementation enables flexibility of input data sizes, this approach significantly increases the inference durations – 4x on 256 × 256 input and 8x on 512 × 512 input, when compared with full input size approaches. Furthermore, the preprocessing step of converting a full-size image into 64 × 64 patches adds an overhead that is directly proportional to the dimensions of the input image. Currently, our denoising models require approximately 3.679 seconds per slice if the input image dimensions are 512 × 512, and 2.575 seconds per slice if the input image dimensions are 256 × 256. Potentially, this could approximately be reduced 0.459 seconds and 0.321 seconds per slice, respectively, if a full input size were instead adopted. The denoising models are also not implemented in an end-to-end workflow – currently, the data needs to be transferred to a designated system via physical storage media. Future work will potentially involve streamlining file I/O to further accelerate DL denoising durations and packaging the pipeline to be tested for deployment at beta site.

#### Explainable AI

While there exist multiple methods that aid in interpretability classification models (Zhou, Khosla and Lapedriza, 2016; Shrikumar, Greenside and Kundaje, 06--11 Aug 2017; Selvaraju, Cogswell and Das, 2017; Smilkov *et al*., 2017), image-to-image model outputs are difficult to explain. Explainable AI techniques such Concept Activation Vectors (CAV) (Clough *et al*., 2019) allow probing the latent spaces of convolutional models to determine which human-friendly concepts the models are most sensitive to. However, this technique can only be applied to network architectures that include a bottleneck layer, such as U-Nets (Ronneberger, Fischer and Brox, 2015), autoencoders (or variants thereof, such as variational-quantized autoencoders (Van Den Oord and Vinyals, 2017)). To the best of our knowledge, there is no prior work on applying CAVs to investigate the performance of image-denoising models. Future work could involve exploring these network architectures to leverage CAVs for explainability.

### B. Conclusion

This work demonstrates an end-to-end framework tailored for AD imaging. The framework involved implementing a LUT to shorten the acquisition duration of an existing brain imaging protocol that was employed at our institution, by sacrificing image quality. Accelerated brain imaging using this faster protocol was demonstrated, and image quality was recovered post-acquisition using DL-based image denoising models. Furthermore, MR physics dictates that the amount of signal captured relates to the volume of the subject being imaged, as this directly affects the size of the proton population. This variability of SNR depending on subject size motivated the authors to implement and demonstrate subject-specific image denoising.

## Supporting information

Supplementary File 3 - AMRI-IP table time

Supplementary File 1 - nnUnet benchmarking

Supplementary File 2 - T1 model selection

## Data Availability

All data produced in the present study are available upon reasonable request to the authors

## V. Acknowledgements

The authors received support from the Biomedical Engineering and Imaging Institute, Dept. of Diagnostic, Molecular and Interventional Radiology, Icahn School of Medicine, Mt. Sinai, New York, NY, USA. Data used in the preparation of this article were obtained from the Alzheimer’s Disease Neuroimaging Initiative (ADNI) database (adni.loni.usc.edu). The ADNI was launched in 2003 as a public-private partnership, led by Principal Investigator Michael W. Weiner, MD. The primary goal of ADNI has been to test whether serial magnetic resonance imaging (MRI), positron emission tomography (PET), other biological markers, and clinical and neuropsychological assessment can be combined to measure the progression of mild cognitive impairment (MCI) and early Alzheimer’s disease (AD). For up-to-date information, see www.adni-info.org. Data collection and sharing for this project was funded by the Alzheimer’s Disease Neuroimaging Initiative (ADNI) (National Institutes of Health Grant U01 AG024904) and DOD ADNI (Department of Defense award number W81XWH-12-2-0012). ADNI is funded by the National Institute on Aging, the National Institute of Biomedical Imaging and Bioengineering, and through generous contributions from the following: AbbVie, Alzheimer’s Association; Alzheimer’s Drug Discovery Foundation; Araclon Biotech; BioClinica, Inc.; Biogen; Bristol-Myers Squibb Company; CereSpir, Inc.; Cogstate; Eisai Inc.; Elan Pharmaceuticals, Inc.; Eli Lilly and Company; EuroImmun; F. Hoffmann-La Roche Ltd and its affiliated company Genentech, Inc.; Fujirebio; GE Healthcare; IXICO Ltd.; Janssen Alzheimer Immunotherapy Research & Development, LLC.; Johnson & Johnson Pharmaceutical Research & Development LLC.; Lumosity; Lundbeck; Merck & Co., Inc.; Meso Scale Diagnostics, LLC.; NeuroRx Research; Neurotrack Technologies; Novartis Pharmaceuticals Corporation; Pfizer Inc.; Piramal Imaging; Servier; Takeda Pharmaceutical Company; and Transition Therapeutics. The Canadian Institutes of Health Research is providing funds to support ADNI clinical sites in Canada. Private sector contributions are facilitated by the Foundation for the National Institutes of Health (www.fnih.org). The grantee organization is the Northern California Institute for Research and Education, and the study is coordinated by the Alzheimer’s Therapeutic Research Institute at the University of Southern California. ADNI data are disseminated by the Laboratory for Neuro Imaging at the University of Southern California.

## References

1. Abadi et al. (2016) ‘TensorFlow: a system for Large-Scale machine learning’, on operating systems … [Preprint]. Available at: https://www.usenix.org/conference/osdi16/technical-sessions/presentation/abadi.

2. Abadi, M. et al. (2016) ‘TensorFlow: Large-Scale Machine Learning on Heterogeneous Distributed Systems’, arXiv [cs.DC]. Available at: http://arxiv.org/abs/1603.04467.

3. Banerjee, D. et al. (2020) ‘Neuroimaging in Dementia: A Brief Review’, Cureus, 12(6), p. e8682.

4. Bateman, R.J. et al. (2012) ‘Clinical and biomarker changes in dominantly inherited Alzheimer’s disease’, The New England journal of medicine, 367(9), pp. 795–804.

5. Block, K.T. (2018) ‘Creating New Value Through Innovation’. ISMRM Workshop on High-Value MRI, 18 February.

6. Clough, J.R. et al. (2019) ‘Global and Local Interpretability for Cardiac MRI Classification’, in Medical Image Computing and Computer Assisted Intervention – MICCAI 2019. Springer International Publishing, pp. 656–664.

7. Commowick, Cervenansky and Cotton (2021) ‘MSSEG-2 challenge proceedings: Multiple sclerosis new lesions segmentation challenge using a data management and processing infrastructure’, MICCAI 2021-24th [Preprint]. Available at: https://hal.inria.fr/hal-03358968/.

8. Deoni, S.C.L. et al. (2022) ‘Simultaneous high-resolution T2 -weighted imaging and quantitative T2 mapping at low magnetic field strengths using a multiple TE and multi-orientation acquisition approach’, Magnetic resonance in medicine: official journal of the Society of Magnetic Resonance in Medicine / Society of Magnetic Resonance in Medicine, 88(3), pp. 1273–1281.

9. Falahati, F. et al. (2015) ‘The use of MRI, CT and lumbar puncture in dementia diagnostics: data from the SveDem Registry’, Dementia and geriatric cognitive disorders, 39(1-2), pp. 81–91.

10. Fedorov, A. et al. (2012) ‘3D Slicer as an image computing platform for the Quantitative Imaging Network’, Magnetic resonance imaging, 30(9), pp. 1323–1341.

11. Geethanath, Poojar and Ravi (2021) ‘MRI denoising using native noise’, Proc Intl Soc Mag [Preprint]. Available at: https://mr.research.columbia.edu/sites/default/files/content/Geethanath%20MRI%20denoising.pdf.

12. Geethanath, S. and Vaughan, J.T., Jr (2019) ‘Accessible magnetic resonance imaging: A review’, Journal of magnetic resonance imaging: JMRI, 49(7), pp. e65–e77.

13. Golshan, H.M., Hasanzadeh, R.P.R. and Yousefzadeh, S.C. (2013) ‘An MRI denoising method using image data redundancy and local SNR estimation’, Magnetic resonance imaging, 31(7), pp. 1206–1217.

14. He et al. (2016) ‘Deep residual learning for image recognition’, Proceedings of the IEEE [Preprint]. Available at: http://openaccess.thecvf.com/content_cvpr_2016/html/He_Deep_Residual_Learning_CVPR_2016_paper.html.

15. Isensee, F. et al. (2021) ‘nnU-Net: a self-configuring method for deep learning-based biomedical image segmentation’, Nature methods, 18(2), pp. 203–211.

16. Jenkinson, M. (2003) ‘Fast, automated, N-dimensional phase-unwrapping algorithm’, Magnetic resonance in medicine: official journal of the Society of Magnetic Resonance in Medicine / Society of Magnetic Resonance in Medicine, 49(1), pp. 193–197.

17. Kingma, D.P. and Ba, J. (2014) ‘Adam: A Method for Stochastic Optimization’, arXiv [cs.LG]. Available at: http://arxiv.org/abs/1412.6980.

18. Loktyushin, A. et al. (2021) ‘MRzero - Automated discovery of MRI sequences using supervised learning’, Magnetic resonance in medicine: official journal of the Society of Magnetic Resonance in Medicine / Society of Magnetic Resonance in Medicine, 86(2), pp. 709–724.

19. Mehan, W.A., Jr et al. (2014) ‘Optimal brain MRI protocol for new neurological complaint’, PloS one, 9(10), p. e110803.

20. Mei, X. et al. (2022) ‘RadImageNet: An Open Radiologic Deep Learning Research Dataset for Effective Transfer Learning’, Radiology: Artificial Intelligence, 4(5), p. e210315.

21. Nair and Hinton (2010) ‘Rectified linear units improve restricted boltzmann machines’, Icml [Preprint]. Available at: https://openreview.net/forum?id=rkb15iZdZB.

22. Patterson, C. (2018) ‘World Alzheimer report 2018’, Alzheimer’s Disease International [Preprint]. Available at: https://apo.org.au/node/260056 (Accessed: 26 September 2022).

23. Pech-Pacheco, J.L. et al. (2000) ‘Diatom autofocusing in brightfield microscopy: a comparative study’, in Proceedings 15th International Conference on Pattern Recognition. ICPR-2000. https://ieeexplore.ieee.org, pp. 314–317 vol.3x.

24. Qian, E. et al. (2022) ‘Tailored magnetic resonance fingerprinting for simultaneous non-synthetic and quantitative imaging: A repeatability study’, Medical physics, 49(3), pp. 1673–1685.

25. Ravi, K.S. et al. (no date) ‘Intelligent Protocolling for Autonomous MRI’, archive.ismrm.org [Preprint]. Available at: https://archive.ismrm.org/2020/4154.html.

26. Ravi, K.S. et al. (no date) ‘MR value driven Autonomous MRI using imr-framework’, in. Joint Annual Meeting ISMRM-ESMRMB 2018.

27. Ronneberger, O., Fischer, P. and Brox, T. (2015) ‘U-Net: Convolutional Networks for Biomedical Image Segmentation’, in Medical Image Computing and Computer-Assisted Intervention – MICCAI 2015. Springer International Publishing, pp. 234–241.

28. Selvaraju, Cogswell and Das (2017) ‘Grad-cam: Visual explanations from deep networks via gradient-based localization’, Proceedings of the Estonian Academy of Sciences. Biology, ecology = Eesti Teaduste Akadeemia toimetised. Bioloogia, okoloogia [Preprint]. Available at: http://openaccess.thecvf.com/content_iccv_2017/html/Selvaraju_Grad-CAM_Visual_Explanations_ICCV_2017_paper.html.

29. Shin, D. et al. (2020) ‘Deep Reinforcement Learning Designed Shinnar-Le Roux RF Pulse Using Root-Flipping: DeepRFSLR’, IEEE transactions on medical imaging, 39(12), pp. 4391–4400.

30. Shrikumar, A., Greenside, P. and Kundaje, A. (06--11 Aug 2017) ‘Learning Important Features Through Propagating Activation Differences’, in D. Precup and Y.W. Teh (eds) Proceedings of the 34th International Conference on Machine Learning. PMLR (Proceedings of Machine Learning Research), pp. 3145–3153.

31. Silva-Spínola, A. et al. (2022) ‘The Road to Personalized Medicine in Alzheimer’s Disease: The Use of Artificial Intelligence’, Biomedicines, 10(2). Available at: https://doi.org/10.3390/biomedicines10020315.

32. Simmons, A. et al. (2011) ‘The AddNeuroMed framework for multi-centre MRI assessment of Alzheimer’s disease: experience from the first 24 months’, International journal of geriatric psychiatry, 26(1), pp. 75–82.

33. Smilkov, D. et al. (2017) ‘SmoothGrad: removing noise by adding noise’, arXiv [cs.LG]. Available at: http://arxiv.org/abs/1706.03825.

34. Snoek, L. et al. (2021) ‘The Amsterdam Open MRI Collection, a set of multimodal MRI datasets for individual difference analyses’, Scientific data, 8(1), p. 85.

35. Thomas, N. et al. (2020) ‘Fully Automated End-to-End Neuroimaging Workflow for Mental Health Screening’, 2020 IEEE 20th International Conference on Bioinformatics and Bioengineering (BIBE) [Preprint]. Available at: https://doi.org/10.1109/bibe50027.2020.00109.

36. Tong, G. et al. (2019) ‘Virtual scanner: MRI on a browser’, Journal of open source software, 4(43), p. 1637.

37. Tong, G., Vaughan, J.T., Jr and Geethanath, S. (2021) ‘Virtual Scanner 2.0: enabling the MR digital twin’, in Proceedings of 2021 ISMRM & SMRT Annual Meeting & Exhibition. 2021 ISMRM & SMRT Annual Meeting & Exhibition.

38. Tsao, J. and Kozerke, S. (2012) ‘MRI temporal acceleration techniques’, Journal of magnetic resonance imaging: JMRI, 36(3), pp. 543–560.

39. Van Den Oord and Vinyals (2017) ‘Neural discrete representation learning’, Advances in neural information processing systems [Preprint]. Available at: https://proceedings.neurips.cc/paper/2017/hash/7a98af17e63a0ac09ce2e96d03992fbc-Abstract.html.

40. Vernooij, M.W. and van Buchem, M.A. (2020) ‘Neuroimaging in Dementia’, in J. Hodler, R.A. Kubik-Huch, and G.K. von Schulthess (eds) Diseases of the Brain, Head and Neck, Spine 2020– 2023: Diagnostic Imaging. Cham (CH): Springer.

41. Walker-Samuel (2019) ‘Using deep reinforcement learning to actively, adaptively and autonomously control of a simulated MRI scanner’, Proc. 27th Annual Meeting of ISMRM [Preprint]. Available at: https://archive.ismrm.org/2019/0473.html.

42. Wang, Z., Simoncelli, E.P. and Bovik, A.C. (2003) ‘Multiscale structural similarity for image quality assessment’, in The Thrity-Seventh Asilomar Conference on Signals, Systems & Computers, 2003. ieeexplore.ieee.org, pp. 1398–1402 Vol.2.

43. Xu, J. et al. (2020) ‘Noisy-As-Clean: Learning Self-supervised Denoising from Corrupted Image’, IEEE transactions on image processing: a publication of the IEEE Signal Processing Society, PP. Available at: https://doi.org/10.1109/TIP.2020.3026622.

44. Zhao, H. et al. (2017) ‘Loss Functions for Image Restoration With Neural Networks’, IEEE Transactions on Computational Imaging, pp. 47–57. Available at: https://doi.org/10.1109/tci.2016.2644865.

45. Zhou, Khosla and Lapedriza (2016) ‘Learning deep features for discriminative localization’, Proceedings of the Estonian Academy of Sciences. Biology, ecology = Eesti Teaduste Akadeemia toimetised. Bioloogia, okoloogia [Preprint]. Available at: http://openaccess.thecvf.com/content_cvpr_2016/html/Zhou_Learning_Deep_Features_CVPR_2016_paper.html.

46. Zhu et al. (2018) ‘AUTOmated pulse SEQuence generation (AUTOSEQ) using Bayesian reinforcement learning in an MRI physics simulation environment’, Proceedings of the 26th [Preprint]. Available at: https://archive.ismrm.org/2018/0438.html.

