## Supplementary Figures for "Accelerated MRI using intelligent protocolling and subject-specific denoising applied to Alzheimer’s disease imaging"

### Slide 1
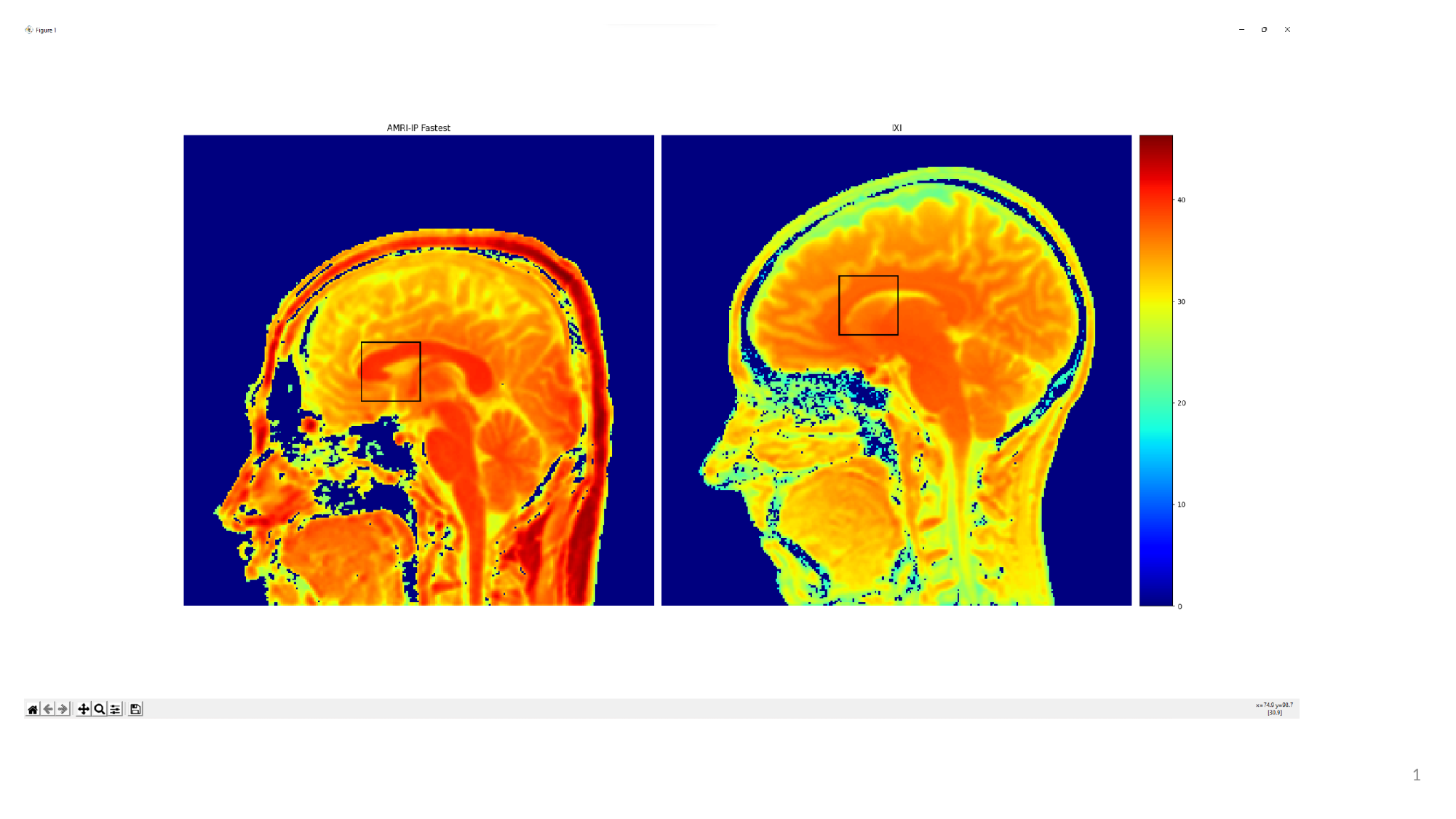

1

### Slide 2
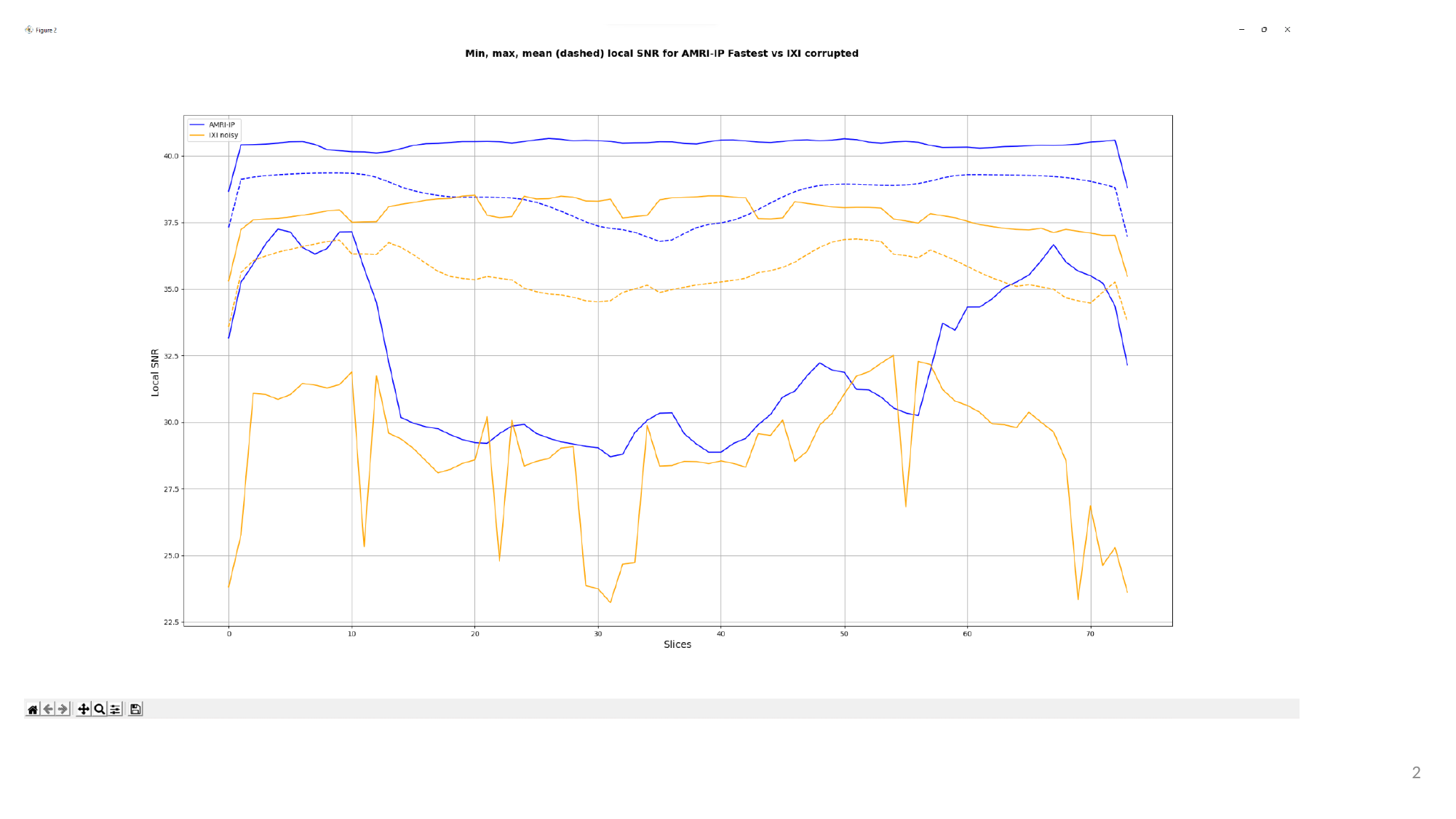

2

### Slide 3
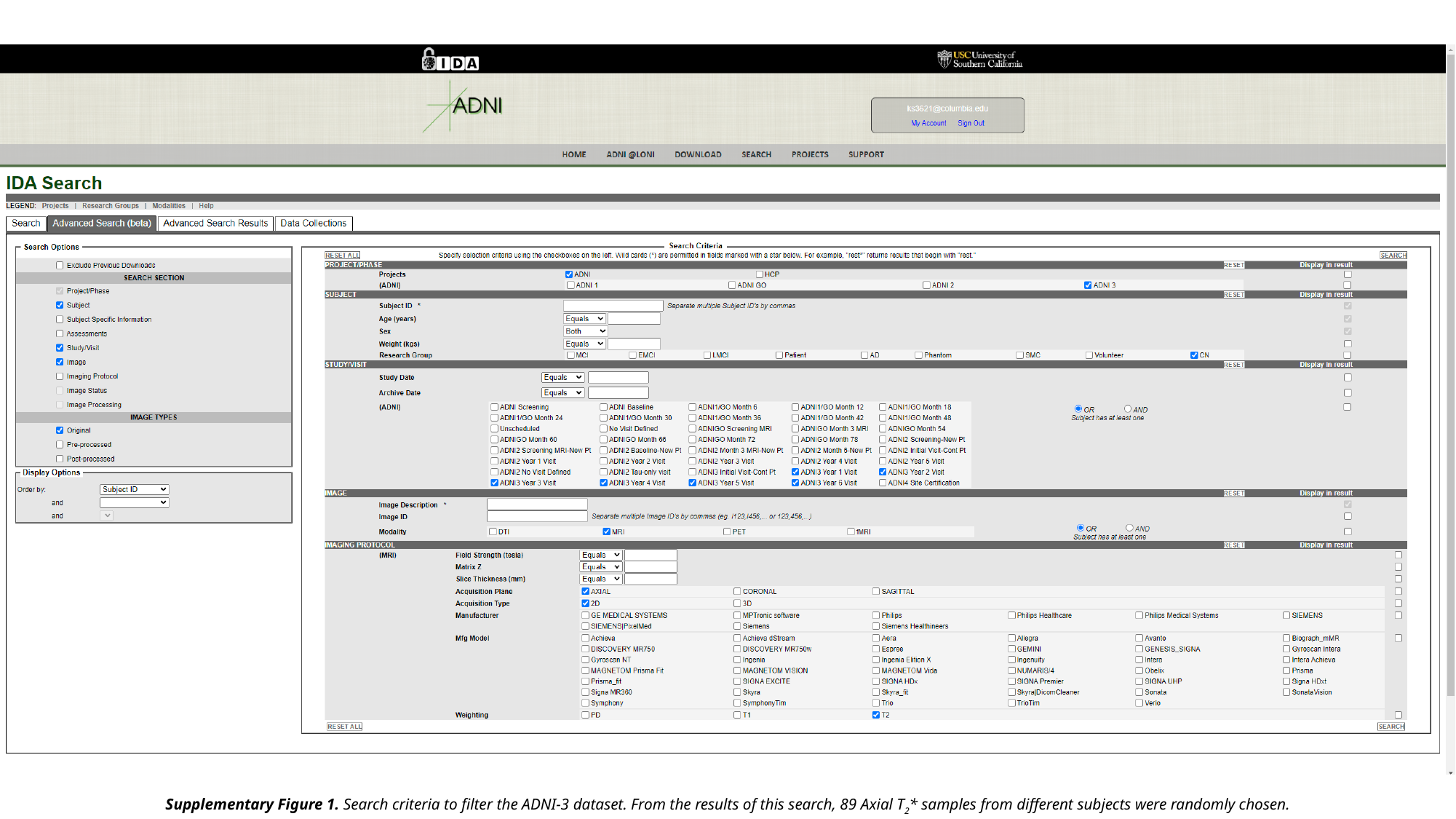

Supplementary Figure 1. Search criteria to filter the ADNI-3 dataset. From the results of this search, 89 Axial T_2_* samples from different subjects were randomly chosen.

### Slide 4
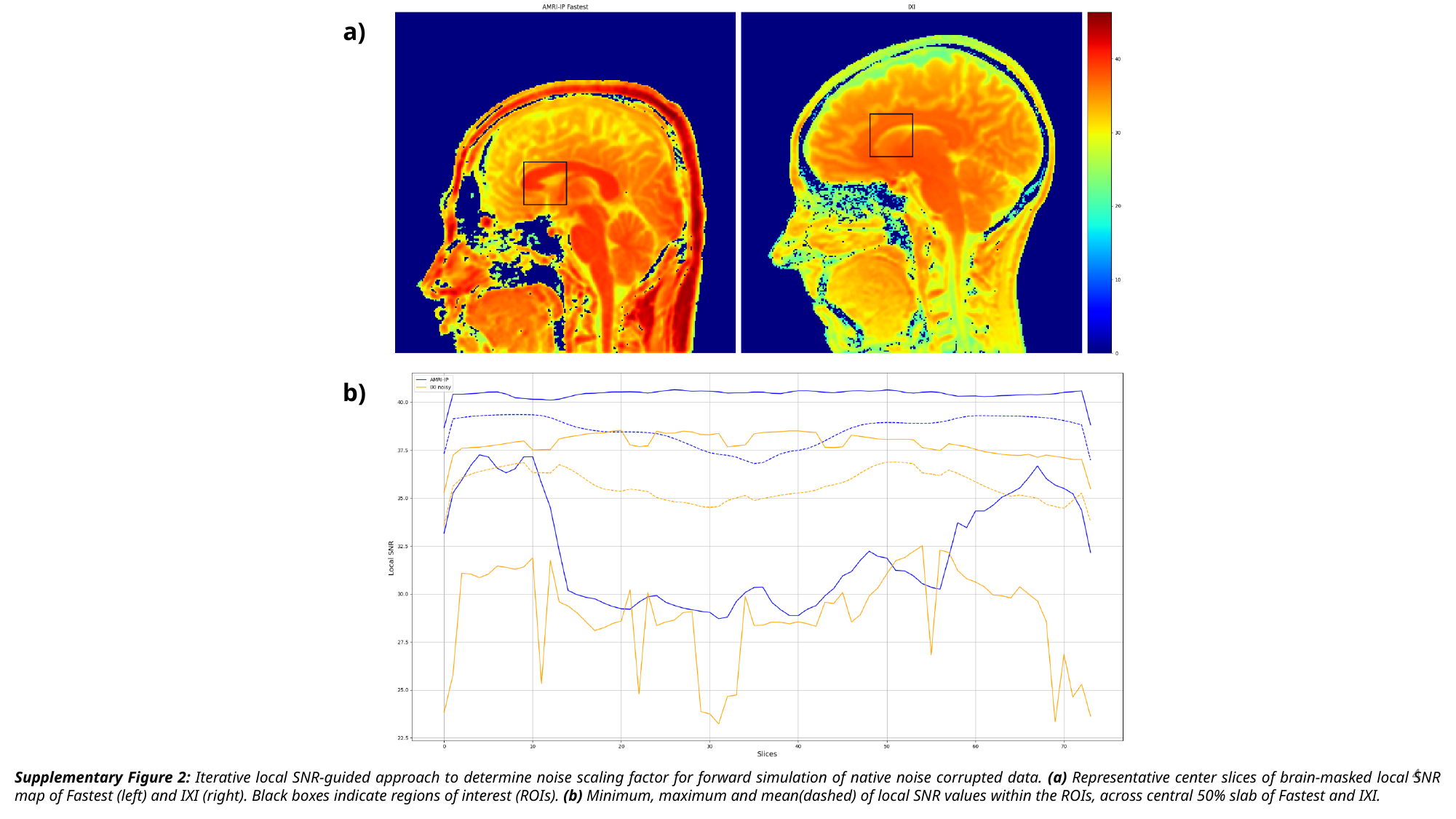

a)
b)
4
Supplementary Figure 2: Iterative local SNR-guided approach to determine noise scaling factor for forward simulation of native noise corrupted data. (a) Representative center slices of brain-masked local SNR map of Fastest (left) and IXI (right). Black boxes indicate regions of interest (ROIs). (b) Minimum, maximum and mean(dashed) of local SNR values within the ROIs, across central 50% slab of Fastest and IXI.

### Slide 5
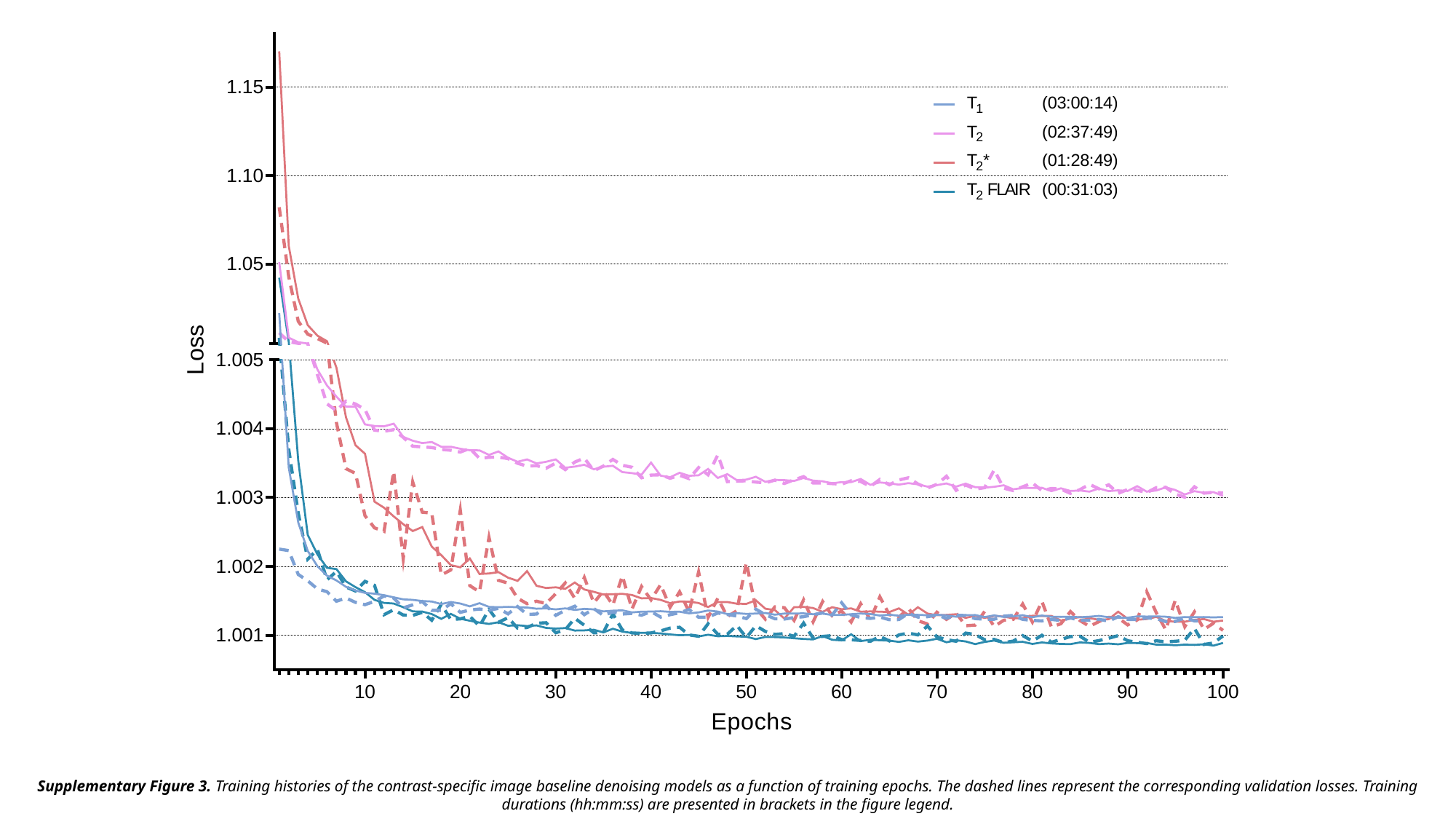

Supplementary Figure 3. Training histories of the contrast-specific image baseline denoising models as a function of training epochs. The dashed lines represent the corresponding validation losses. Training durations (hh:mm:ss) are presented in brackets in the figure legend.

### Slide 6
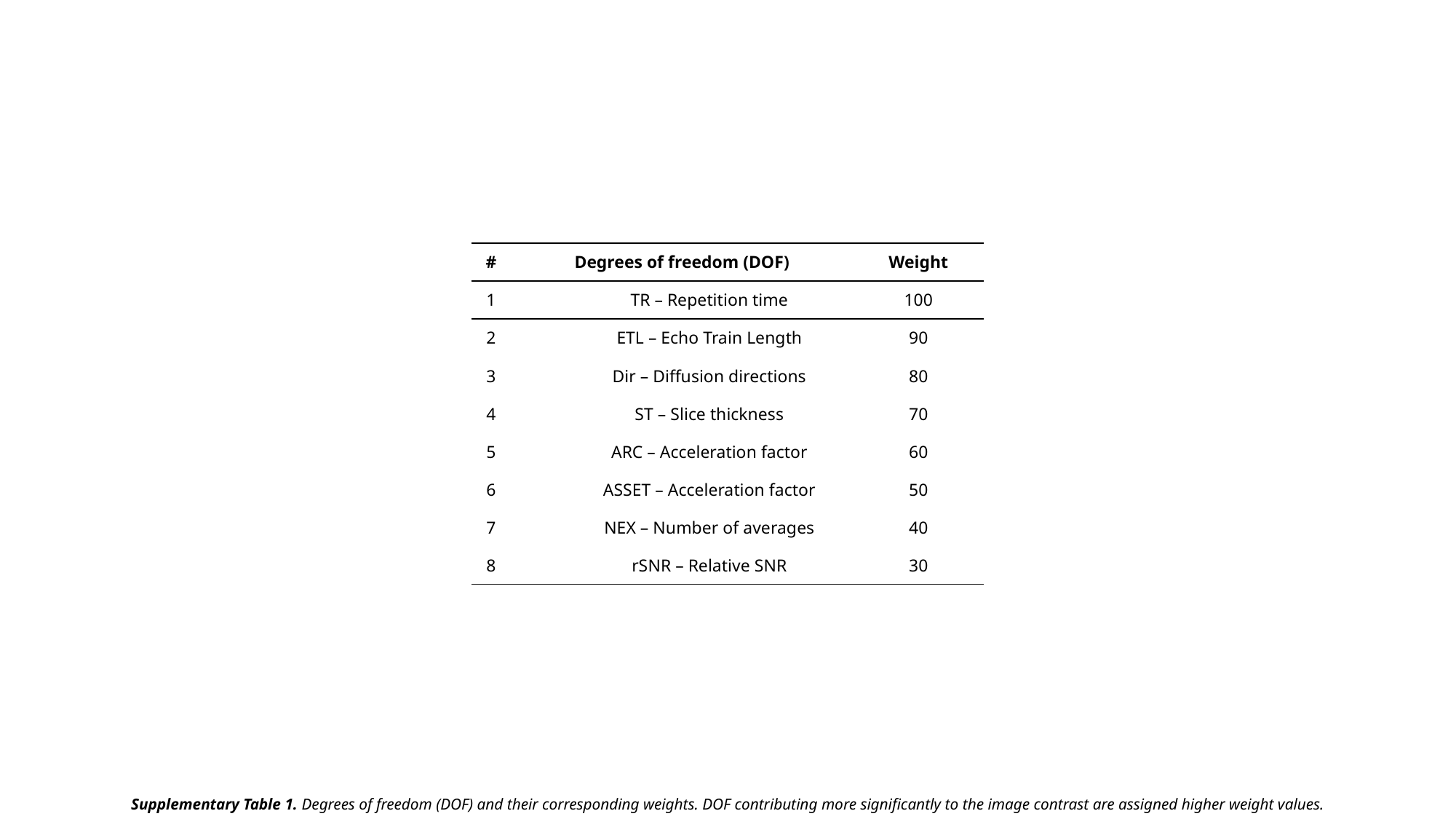

| # | Degrees of freedom (DOF) | Weight |
| --- | --- | --- |
| 1 | TR – Repetition time | 100 |
| 2 | ETL – Echo Train Length | 90 |
| 3 | Dir – Diffusion directions | 80 |
| 4 | ST – Slice thickness | 70 |
| 5 | ARC – Acceleration factor | 60 |
| 6 | ASSET – Acceleration factor | 50 |
| 7 | NEX – Number of averages | 40 |
| 8 | rSNR – Relative SNR | 30 |
Supplementary Table 1. Degrees of freedom (DOF) and their corresponding weights. DOF contributing more significantly to the image contrast are assigned higher weight values.

### Slide 7
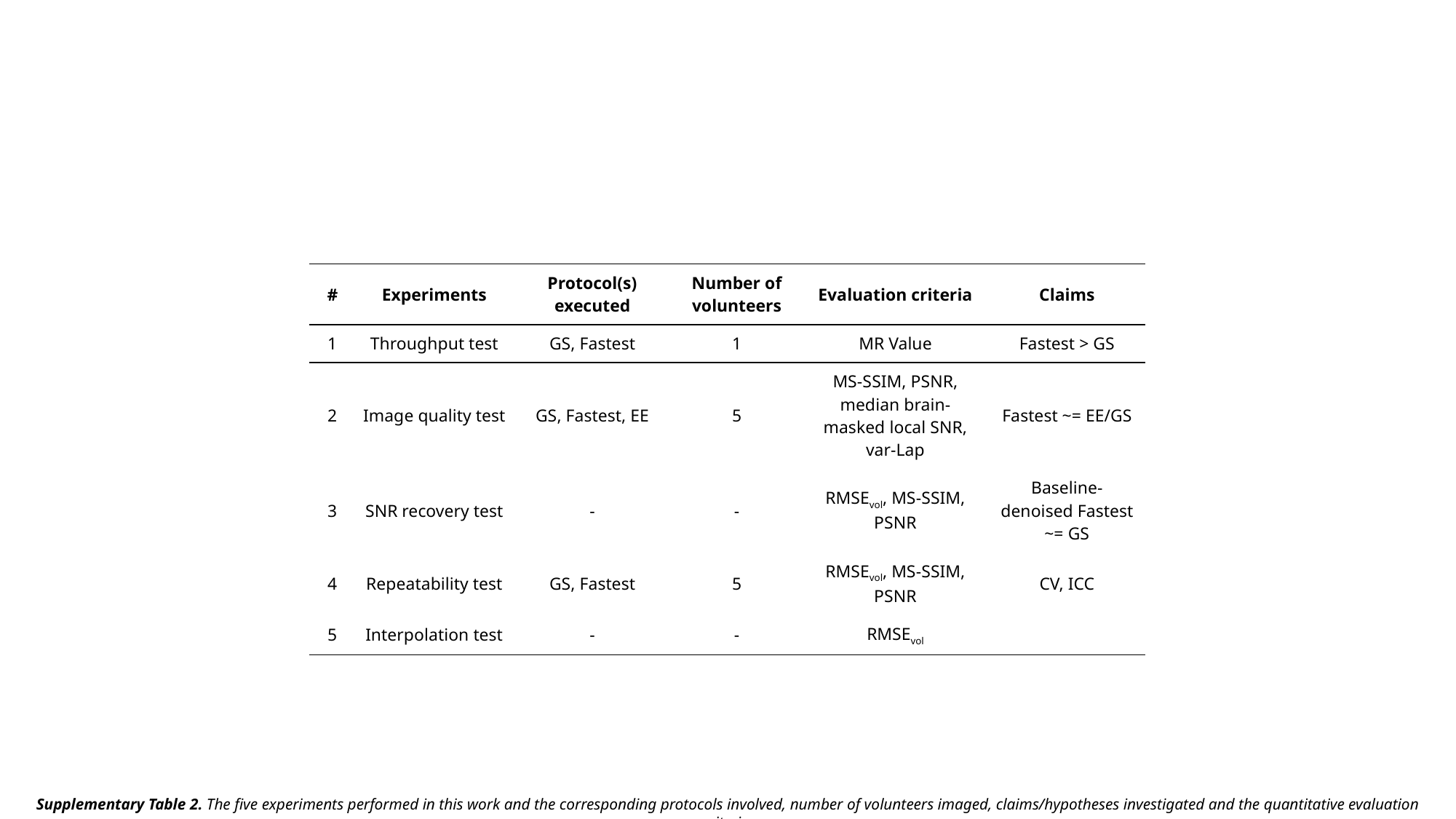

| # | Experiments | Protocol(s) executed | Number of volunteers | Evaluation criteria | Claims |
| --- | --- | --- | --- | --- | --- |
| 1 | Throughput test | GS, Fastest | 1 | MR Value | Fastest > GS |
| 2 | Image quality test | GS, Fastest, EE | 5 | MS-SSIM, PSNR, median brain-masked local SNR, var-Lap | Fastest ~= EE/GS |
| 3 | SNR recovery test | - | - | RMSE_vol_, MS-SSIM, PSNR | Baseline-denoised Fastest ~= GS |
| 4 | Repeatability test | GS, Fastest | 5 | RMSE_vol_, MS-SSIM, PSNR | CV, ICC |
| 5 | Interpolation test | - | - | RMSE_vol_ | |
Supplementary Table 2. The five experiments performed in this work and the corresponding protocols involved, number of volunteers imaged, claims/hypotheses investigated and the quantitative evaluation criteria.
